# A Randomised Clinical Trial of the Safety and Pharmacokinetics of VRC07-523LS Administered via Different Routes and Doses (HVTN 127/HPTN 087)

**DOI:** 10.1101/2024.01.10.23299799

**Authors:** Stephen R. Walsh, Cynthia L. Gay, Shelly T. Karuna, Ollivier Hyrien, Timothy Skalland, Kenneth H. Mayer, Magdalena E. Sobieszczyk, Lindsey R. Baden, Paul A. Goepfert, Carlos del Rio, Guiseppe Pantaleo, Philip Andrew, Carissa Karg, Zonglin He, Helen Lu, Carmen A. Paez, Jane A. G. Baumblatt, Laura L. Polakowski, Wairimu Chege, Sophie Janto, Xue Han, Yunda Huang, Julie Dumond, Margaret E. Ackerman, Adrian B. McDermott, Britta Flach, Estelle Piwowar-Manning, Kelly Seaton, Georgia D. Tomaras, David C. Montefiori, Lucio Gama, John R. Mascola, HVTN 127/HPTN 087 Study Team

**Affiliations:** Division of Infectious Diseases, Brigham and Women’s Hospital, Boston, MA; Harvard Medical School, Boston, MA; Division of Infectious Diseases, University of North Carolina, Chapel Hill, NC; Vaccine and Infectious Disease Division, Fred Hutchinson Cancer Center, Seattle, WA; Fenway Institute, Boston, MA; Division of Infectious Diseases, Columbia University, New York, NY; University of Alabama at Birmingham, Birmingham, AL; Emory University School of Medicine and Ponce de Leon Center of the Grady Health System, Atlanta, GA; Centre Hospitalier Universitaire Vaudois, Lausanne, Switzerland; FHI 360, Durham, NC; Division of AIDS, National Institute of Allergy and Infectious Diseases, Bethesda, MD; Eshelman School of Pharmacy, University of North Carolina, Chapel Hill, NC; Thayer School of Engineering, Dartmouth College, Hanover, NH; Johns Hopkins University, Baltimore, MD; Department of Surgery and Duke Human Vaccine Institute, Duke University, Durham, NC; Vaccine Research Center, National Institute of Allergy and Infectious Diseases, Bethesda, MD

## Abstract

**Background:** Broadly neutralizing antibodies (bnAbs) are a promising approach for HIV-1 prevention. In the only bnAb HIV prevention efficacy studies to date, the Antibody Mediated Prevention (AMP) trials, a CD4-binding site targeting bnAb, VRC01, administered intravenously (IV), demonstrated 75% prevention efficacy against highly neutralization-sensitive viruses but was ineffective against less sensitive viruses. Greater efficacy is required before passively administered bnAbs become a viable option for HIV prevention; furthermore subcutaneous (SC) or intramuscular (IM) administration may be preferred. VRC07-523LS is a next-generation bnAb targeting the CD4-binding site and was engineered for increased neutralization breadth and half-life.

**Methods:** Participants were recruited between 02 February 2018 and 09 October 2018. 124 healthy participants without HIV were randomized to receive five VRC07-523LS administrations via IV (T1: 2.5 mg/kg, T2: 5 mg/kg, T3: 20 mg/kg), SC (T4: 2.5 mg/kg, T5: 5 mg/kg) or IM (T6: 2.5 mg/kg or P6: placebo) routes at four-month intervals. Safety data were collected for 144 weeks following the first administration. VRC07-523LS serum concentrations were measured by ELISA after the first dose through Day 112 in all participants and by binding antibody multiplex assay (BAMA) thereafter in 60 participants (10 per treatment group) through Day 784. Compartmental population pharmacokinetic (PK) analyses were conducted to evaluate the VRC07-523LS serum pharmacokinetics. Neutralization activity was measured in a TZM-bl assay and anti-drug antibodies (ADA) were assayed using a tiered bridging assay testing strategy.

**Results:** Injections were well-tolerated, with mild pain or tenderness reported commonly in the SC and IM groups, and mild to moderate erythema or induration reported commonly in the SC groups. Infusions were generally well-tolerated, with infusion reactions reported in 3 of 20 participants in the 20 mg/kg IV group. Peak geometric mean (GM) concentrations (95% confidence intervals) following the first administration were 29.0 μg/mL (25.2, 33.4), 58.5 μg/mL (49.4, 69.3), and 257.2 μg/mL (127.5, 518.9) in T1-T3 with IV dosing; 10.8 μg/mL (8.8, 13.3) and 22.8 μg/mL (20.1, 25.9) in T4-T5 with SC dosing; and 16.4 μg/mL (14.7, 18.2) in T6 with IM dosing. Trough GM concentrations immediately prior to the second administration were 3.4 μg/mL (2.5, 4.6), 6.5 μg/mL (5.6, 7.5), and 27.2 μg/mL (23.9, 31.0) with IV dosing; 0.97 μg/mL (0.65, 1.4) and 3.1 μg/mL (2.2, 4.3) with SC dosing, and 2.6 μg/mL (2.05, 3.31) with IM dosing. Peak VRC07-523LS serum concentrations increased linearly with the administered dose. At a given dose, peak and trough concentrations, as well as serum neutralization titres, were highest in the IV groups, reflecting the lower bioavailability following SC and IM administration. A single participant was found to have low titre ADA at a lone timepoint. VRC07-523LS has an estimated mean half-life of 42 days (95% CI: 40.5, 43.5), approximately twice as long as VRC01.

**Conclusions:** VRC07-523LS was safe and well-tolerated across a range of doses and routes and is a promising long-acting bnAb for inclusion in HIV-1 prevention regimens.

## Introduction

Despite the proven effectiveness of behavioural and pharmacological methods of preventing HIV-1 (1–3), worldwide over 1.5 million individuals are estimated to acquire HIV yearly (4). Accordingly, long-acting and well-tolerated biomedical prevention approaches such as passive immunization using broadly neutralizing antibodies (bnAbs) are a promising new approach for HIV-1 prevention (5, 6). In the only efficacy trials of a bnAb against HIV-1 conducted to date, the Antibody Mediated Prevention (AMP) trials, overall efficacy was limited (7). Importantly however, the AMP studies provided proof of concept that passive immunization with a bnAb could confer protection against HIV-1 infection and identified neutralization as a correlate of protection (8). Despite overall low efficacy, viruses that were highly sensitive to neutralization by VRC01 (IC_80_ <1 μg/mL) were successfully blocked in vivo (7, 8). This result suggests that bnAbs with increased breadth and increased potency might provide increased protection.

Toward this aim, another antibody (VRC07) was identified from the same donor as VRC01, cloned, and then engineered to increase its neutralization potency and breadth (7). Additional mutations (Met428Leu and Asn434Ser;-LS) were made to the Fc portion of the antibody to increase the antibody’s binding affinity for the neonatal Fc receptor (FcRn), resulting in increased recirculation and therefore a longer half-life in vivo (8, 9). The resultant antibody, VRC07-523LS, was 5-to 8-fold more potent than VRC01 and considerably broader in vitro, with an IC_50_ < 1 μg/mL against 92% of HIV-1 pseudoviruses across circulating HIV-1 clades (7). In a first-in-human, dose-escalation study (VRC 605), VRC07-523LS administered intravenously (IV) to healthy volunteers without HIV was safe, well-tolerated, and found to have a half-life of 38 days, more than twice the half-life of VRC01 (10).

The increased potency and durability of VRC07-523LS relative to VRC01 may help address a key question regarding bnAbs as a potential HIV prevention tool, namely the feasibility of their administration. More potent and longer-acting bnAbs can be administered less frequently and at lower doses and thus at lower cost. The route of administration is another important feasibility consideration, influenced by both dose and volume. In the AMP studies, the necessary dose and volume of the less potent VRC01 required IV administration. While IV administration was well-tolerated in the AMP studies, the World Health Organization (WHO)’s 2022 target product profile for HIV prevention bnAbs reflects a preference for subcutaneous (SC) or intramuscular (IM) injection, particularly for broad programmatic deployment in low and middle income countries (LMICs) (11).

In planning for an eventual efficacy trial with combination bnAb administration, we conducted a phase 1 study to comprehensively assess the safety and pharmacokinetics of VRC07-523LS at a range of doses and via several different routes of administration.

## Methods

### Participants and Study Design

HVTN 127/HPTN 087 is a multicenter, randomized, partially blinded phase 1 clinical trial to evaluate the safety and serum concentrations of VRC07-523LS, administered in multiple doses and routes to healthy adults without HIV (**Table 1**). Participants received five administrations of the assigned dose and route every four months at Weeks 0, 16, 32, 48, and 64 and were followed on study through Week 112. The product was vialed at a concentration of 100 mg/mL and volume considerations limited the SC route to a maximum of 5 mg/kg and the IM route to 2.5 mg/kg.

**Table 1.** Study schema.

| Group | N | Dose | Route | Month<br>0 | Month<br>4 | Month<br>8 | Month<br>12 | Month<br>16 |
| --- | --- | --- | --- | --- | --- | --- | --- | --- |
| T1 | 19 | 2.5 mg/kg | IV | X | X | X | X | X |
| T2 | 19 | 5 mg/kg | IV | X | X | X | X | X |
| T3 | 21 | 20 mg/kg | IV | X | X | X | X | X |
| T4 | 21 | 2.5 mg/kg | SC | X | X | X | X | X |
| T5 | 20 | 5 mg/kg | SC | X | X | X | X | X |
| T6 | 21 | 2.5 mg/kg | IM | X | X | X | X | X |
| P6 | 3 | Placebo | IM | X | X | X | X | X |

IV – intravenous; SC – subcutaneous; IM – intramuscular

Participants were enrolled at seven clinical research sites (CRSs): Brigham & Women’s Hospital (Boston, MA), the Fenway Institute (Boston, MA), the University of North Carolina (Chapel Hill, NC), the University of Alabama (Birmingham, AL), Ponce de Leon CRS (Atlanta, GA), Columbia University (New York, NY), and the Centre Hospitalier Universitaire Vaudois (Lausanne, Switzerland). Volunteers were eligible if they were between 18 and 50 years of age, without HIV-1, in good overall health, and unlikely to acquire HIV during the study period based on behaviours reported within the 12 months prior to enrollment (12). The study was approved by the Institutional Review Boards of each participating CRS and all participants provided written informed consent. The HIV Vaccine Trials Network (HVTN) Safety Monitoring Board (SMB) provided safety oversight. The trial was registered with ClinicalTrials.gov (NCT03387150).

Participants were randomized through an Internet-based randomization system in blocks. Group 6 was added via an amendment after enrollment of Groups 1 to 5 began. Participants and study staff were unblinded to group assignment (Groups 1 through 6) but blinded to antibody versus placebo within Group 6.

### Safety Assessments

Following the first study product administration (SPA), participants were monitored for at least 60 minutes to assess for solicited local and systemic adverse events (AEs), including pain/tenderness at the infusion or injection site, fever, malaise, myalgia, headache, chills, arthralgia, nausea, urticaria, non-exertional dyspnea, non-exertional tachycardia, generalized pruritus, facial flushing, and unexplained diaphoresis. If no reactions occurred after the first administration, in-clinic monitoring after subsequent SPAs could decrease to 30 minutes. Participants completed daily symptom diaries to document solicited AEs for three days following each infusion or injection. Symptom-targeted physical exams and laboratory assays (complete blood count with differential, creatinine, alanine aminotransferase, urine dipstick, pregnancy testing, and HIV testing) were performed for safety monitoring at prespecified intervals throughout the study. Unsolicited AEs were collected throughout the study.

### Pharmacokinetics Studies

#### Enzyme Linked Immunosorbent Assay (ELISA)

An ELISA assay was used to measure serum concentrations of VRC07-523LS between the first and second SPA. These concentrations were quantified in 96 well plates on a Beckman Biomek based automation platform. The monoclonal antibody (mAb) 5C9 was coated onto Immulon-4HXB microtitre plates overnight at 4°C. Plates were then washed and blocked (10% FBS in PBS) for 2 hours at room temperature. Duplicate serial 3-fold dilutions covering the range of 1:100 – 1:24 300 of the test sample were incubated 2 hours at 37°C followed by Horseradish Peroxidase (HRP) – labeled goat anti-human antibody (1 hour at 37°C) and TMB substrate (15 minutes at room temperature). Color development was stopped by addition of sulfuric acid and plates were read within 30 minutes at 450 nm via the Molecular Devices Paradigm plate reader. Linear regression of a standard curve of VRC07-523LS covering the range from 0.91 to 5 ng/mL was utilized to quantitate sample concentrations based upon the average of sample dilutions within the range of the assay. The lower limit of quantification (LLoQ) of the ELISA assay was 1 μg/mL. Serum concentration data from 121 volunteers were measured by the ELISA assay on Target Visit Days 0, 3, 6, 28, 56, 84, and 112 (second SPA).

#### Binding Antibody Multiplex Array (BAMA)

For programmatic reasons, the BAMA assay was used to measure serum concentrations of VRC07-523LS in samples collected after the second SPA in a subset of participants enrolled in each treatment group. Serum VRC07-523LS concentrations were measured on a Bio-Plex instrument (Bio-Rad) using a validated assay designed to measure infused VRC07-523LS by its ability to bind anti-idiotype antibody captured on fluorescent magnetic beads. This assay was derived from a standardized custom HIV-1 Luminex assay (13–16). The Bioplex software provides 2 readouts: a background-subtracted median fluorescent intensity (MFI), where background refers to a plate level control (i.e., a blank well containing antigen-conjugated beads run on each plate), and a concentration based on a standard curve using a 5PL curve fit. Each sample was run in duplicate.

VRC07-523LS was titrated and combined to create a standard curve used to determine concentration of the diluted samples. The negative controls were CH58 (an irrelevant mAb) and blank beads. Samples with VRC07-523LS concentrations below 0.01 μg/mL at a dilution of 1:100 were truncated at 0.01 μg/mL for plotting purposes. All samples are shown in the plot, including concentrations below the LLoQ. Samples with concentrations above the LLoQ at a 1:100 dilution were further tested at various dilution factors to obtain MFIs in the linear range of the standard curve, and the in-well concentration closest to the EC_50_ of the 5PL standard curve was reported.

Several criteria were used to determine if data from an assay were acceptable and could be statistically analyzed. The standard curve EC_50_ values and MFI values were tracked against historical data in Levey Jennings and points with an MFI > 100 must have had a %CV <20% between replicates. Any sample without at least 2 observed concentrations in agreement with each other, or with baseline MFI > 1000 was repeated to obtain an accurate measurement. The physiological LLoQ of the BAMA assay was 0.0457 μg/mL. Serum concentrations were measured using BAMA in 60 of 121 participants who received the study product (10 randomly selected participants for each treatment group) in samples collected at target visit days 168, 224, 280, 336, 392, 448, 504, 560, 616, 672, 728, 784.

#### Neutralization activity

Neutralizing antibody activity against HIV-1 was measured as a function of reduction in Tat-regulated luciferase (Luc) reporter gene expression in TZM-bl cells as described (17, 18). The assay performed in TZM-bl cells measured neutralization titres against Env-pseudotyped viruses sensitive to the bnAb (i.e., bnAb-specific viruses) in each group and a magnitude-breadth panel of Env-pseudotyped viruses. The assay tested neutralization of a panel of Tier 2 viruses that exhibit a range of known sensitivities to VRC07-523LS (**Supplemental Table 1**), which are H703_0646_051sN, H703_1471_190s, H703_1750_140Es, H704_0726_080sN, H704_1535_030sN, H704_2544_140eN01, PVO.4 (19), and a negative control Env-pseudotyped virus (SVA-MLV). Serum neutralization titre was defined as the serum dilution that reduced relative luminescence units (RLU) by 50% and 80% (ID_50_ and ID_80_) relative to the RLU in virus control wells (cells + virus only) after subtraction of background RLU (cells only).

### Anti-Drug Antibodies

Anti-drug antibodies (ADA) were assayed and characterized using a tiered bridging assay testing strategy, in which ADAs act to bridge labeled drug product and are detected in an electrochemiluminescent assay, as previously described (20). In Tier I, a sensitive binding assay determined if samples may have ADA present. In Tier II, the response was confirmed by establishing the specificity of the response by competition with free drug. In Tier III, the ADA response magnitude was characterized by titration.

### Statistical Methods

#### Endpoints

The first primary endpoint was the safety and tolerability of different doses of VRC07-523LS administered IV, SC, and IM by repeat dosing every 16 weeks for a total of 5 administrations. The second primary endpoint was the serum concentrations of VRC07-523LS administered IV, SC, and IM over 6 different dose/route regimens. The safety data analyses included data from all enrolled participants; the concentration data analyses included data from 121 participants who received VRC07-523LS (data from the three placebo recipients were excluded).

#### Statistical approaches

Magnitude-breadth (MB) curves (21) were constructed by study week for ID_50_ and ID_80_ separately. All statistical analyses were carried out using R version 4.0.

#### Population Pharmacokinetics Analysis

The population pharmacokinetic (PK) profile of VRC07-523LS in serum was described using compartmental models. The analysis considered and compared PK models with one, two, and three compartments. These candidate models were fitted to observed serum concentrations using a nonlinear mixed effects approach, including random effects to account for inter-participant variability in PK parameters. The conditional variance of the error term, given the random effects, was assumed to include two additive terms: one term was proportional to the conditional expectation of the concentration; the second term was a constant. Participant-specific (individual) PK parameters were assumed to follow log-normal distributions. The variance-covariance matrix of the random effects was fully unstructured. Additionally, PK parameters were adjusted for the assay (ELISA or BAMA) used to measure concentrations. The model was fitted using the method of maximum likelihood, implemented using the Stochastic Approximation Expectation Maximization (SAEM) algorithm. The significance of the difference in each PK parameter between the two assays was evaluated using Wald tests. Comparison of models with one, two, and three compartments was carried out using a corrected Bayes Information Criterion (BICc). Participant-specific PK parameters were predicted using their empirical Bayes estimates, defined as the most probable value of the participant-specific PK parameters, given the estimated population parameters and the data from the corresponding participant. These participant-specific PK parameters were used to predict the most probable trajectory of the serum concentration for each participant. Modeling assumptions were evaluated using weighted conditional residuals plotted as a function of study day. Model-based predicted participant-specific (most probable) serum concentration trajectories were plotted as a function of study day and compared to the observed serum concentrations. All compartmental PK analyses were conducted using MonolixSuite Version 2019R2 (22).

The area under the curve (AUC) of the VRC07-523LS serum concentration of each individual study participant after one study product administration was computed by summing the estimate of the AUC between Days 0 and 112 computed using the trapezoidal method applied to untransformed serum concentrations and the estimate of the AUC after Day 112 computed as exp(a+112*b)/exp(b) where a and b are the intercept and slope of a linear regression model describing log-serum concentration as a function of time since baseline and fitted to serum concentrations measured in the log-linear portion of the pharmacokinetics of VRC07-523LS.

#### Neutralization Activity

Key secondary objectives included determining whether serum neutralizing activity is maintained at consistent levels after each product administration and to determine whether serum neutralizing activity agreed with measured serum concentrations. A response to a given pseudovirus was considered positive if the corresponding ID_50_ or ID_80_ value was > 10 or 20, depending on the starting dilution of the sample. The assay measured neutralisation titres against seven Env-pseudotyped viruses to determine bnAbs activities against specific viruses (**Supplemental Table 1**).

## Results

### Participant Characteristics and Demographics

Participants were recruited between 02 February 2018 and 09 October 2018. A total of 124 participants enrolled between 28 February 2018 and 09 October 2018 at six sites in the US and one in Switzerland (**Figure 1**). 75 participants (60%) were assigned female sex at birth; seven participants reported that they were gender non-conforming. 71 participants (57%) identified as White, 28 (23%) as Black/African American, and 12 (10%) as Hispanic (**Table 2**). The median age of participants at enrollment was 28 years. Ninety-five participants (77%) received all five injections or infusions. The final SPA occurred on 10 January 2020, prior to the arrival of COVID-19 in the US or Switzerland, however pandemic restrictions had a considerable impact on follow-up visits. To ensure that sites could collect safety information, remote visits were permitted to be conducted, but no samples could be collected until local safety restrictions allowed in-person visits. The final study visit occurred on 04 December 2020. Sample availability is shown in **Figure 1**.

**Figure 1.**
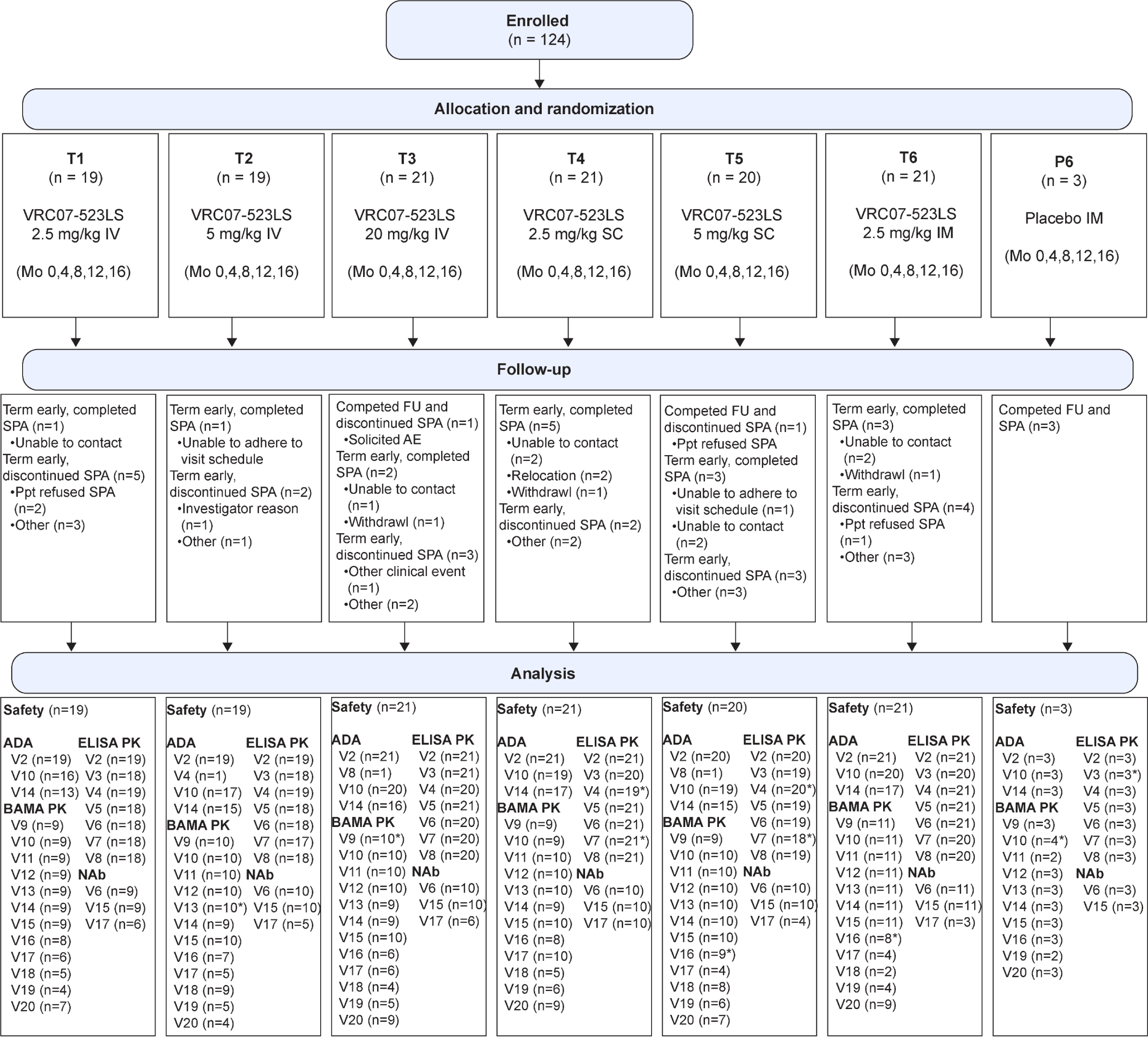
CONSORT diagram illustrating participant flow through the study.

**Table 2.** Participant demographics.

|  | **Female** | **Male** | **Total** |
| --- | --- | --- | --- |
| Black or African-American | 19 | 9 | 28 (23%) |
| Asian | 3 | 3 | 6 (5%) |
| White | 41 | 30 | 71 (57%) |
| Other | 6 | 2 | 8 (7%) |
| Multi-racial | 6 | 5 | 11 (9%) |
| Hispanic | 7 | 5 | 12 (10%) |
| Non-Hispanic | 68 | 44 | 112 (90%) |
| **Total** | **75 (60%)** | **49 (40%)** | **124** |

### Safety and Tolerability

VRC07-523LS was safe and generally well-tolerated at all doses and via all routes administered (**Figure 2**). Mild to moderate injection site pain and/or tenderness was noted by most participants randomized to receive VRC07-523LS by the SC or IM routes. Erythema and/or induration at the injection site was noted by most participants in the SC groups but was generally mild to moderate. Malaise/fatigue and headaches were the most common systemic solicited AEs.

**Figure 2.**
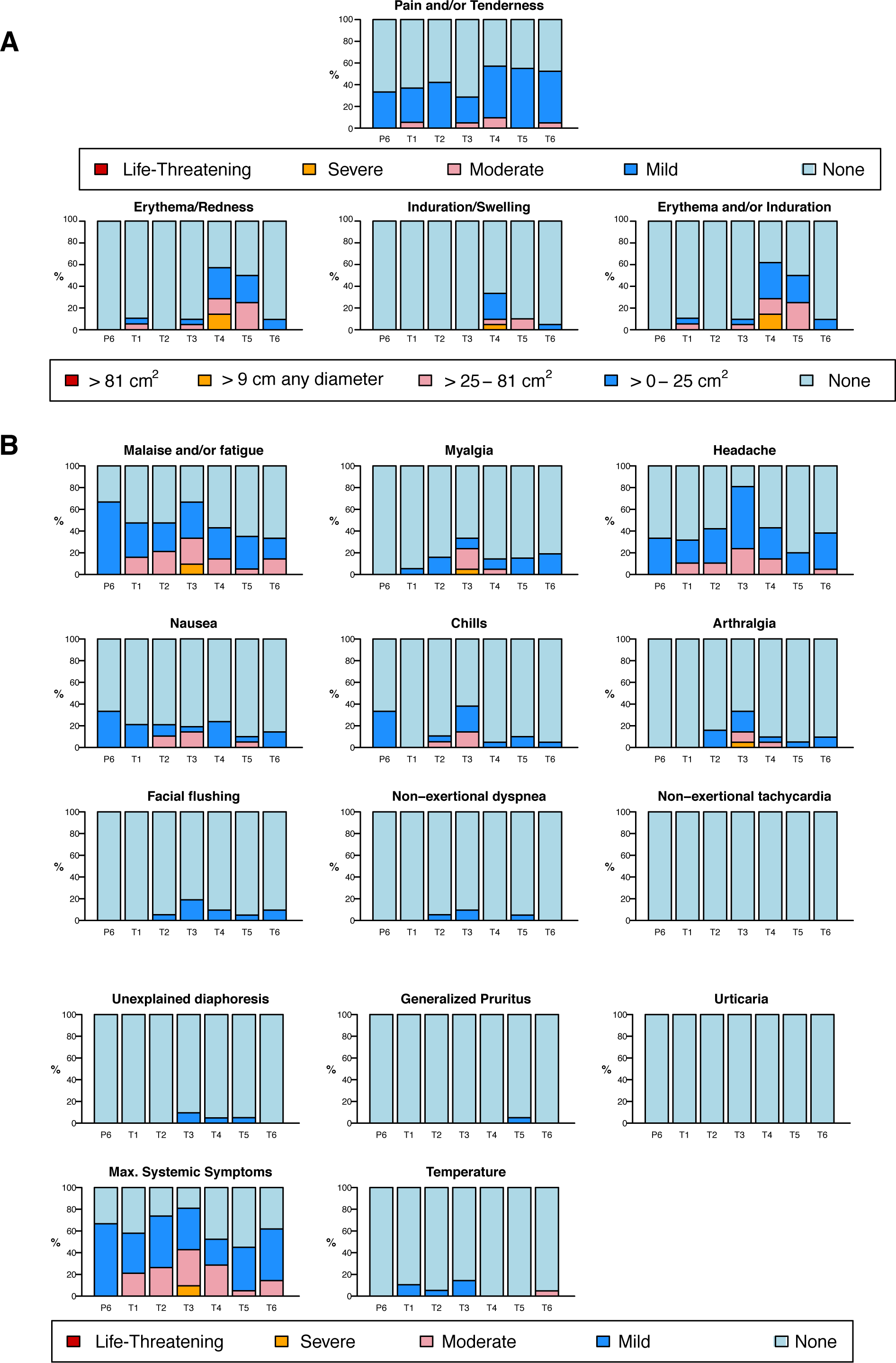
Solicited adverse events (AEs) reported by participants after product administration. Local (A) and systemic (B) solicited AEs were collected for three days following each administration of VRC07-523LS or placebo. P6 = placebo IM; T1 = 2.5 mg/kg IV; T2 = 5 mg/kg IV; T3 = 20 mg/kg IV; T4 = 2.5 mg/kg SC; T5 = 5 mg/kg SC; T6 = 2.5 mg/kg IM.

The majority (98 [79%]) of participants experienced at least one AE during the study, but the vast majority were deemed unrelated to VRC07-523LS administration. Of the sixteen unsolicited AEs deemed related to VRC07-523LS, four began as solicited AEs that continued past the diary period and nine (7% of participants) were isolated symptoms or combinations of symptoms suggestive of infusion-related reactions, similar to those seen with VRC01 administration (23). These symptoms included generalized pruritis, facial flushing, chills, myalgia, arthralgia, nausea, headache, and diaphoresis.

In three participants in the 20 mg/kg IV group, these combinations of symptoms were deemed to be consistent with infusion reactions. No pregnancies or HIV acquisitions were reported during the study. There were three serious adverse events (SAEs) all deemed unrelated to VRC07-523LS: *Staphylococcal* infection, pyrexia in a returned traveller, and urinary calculus.

Participants completed acceptability surveys at every SPA visit to assess their opinions on elements of the administration including discomfort, overall time commitment, and willingness to receive products via their specific route (**Supplemental Table 2**). In general, participants reported that the discomfort, pain, and anxiety related to the SPA was acceptable, although participants in the two SC groups were more likely to say that these were unacceptable (**Supplemental Tables 2A-2C**). The time required for SPA was considered acceptable by most participants (>87% across all visits and groups; **Supplemental Table 2D**). Most participants were very or somewhat willing to use the same method to prevent a serious illness (**Supplemental Table 2E**) and most would recommend that method to a friend at risk of HIV, but this recommendation was lowest for participants in the SC groups at the final SPA (77.4% yes; **Supplemental Table 2F).** Four participants reported adverse social impacts (12) as a result of their participation, mostly in the realm of their interpersonal relationships.

### Anti-drug Antibody

The presence and magnitude of ADA in serum were measured using a tiered “bridging assay” to detect ADA from serum samples obtained at Months 0, 2, 4, 8, and 16. A total of 124 participant samples were tested from Month 0, 1 from Month 2, 2 from Month 4, 114 from Month 8 and 96 from Month 16. A single participant enrolled in Group 4 (SC route, 2.5 mg/kg VRC07-523LS) exhibited a confirmed ADA response at Month 8 (4 months after the 2^nd^ SPA) with a low titre of 1:9 but was ADA negative at Month 16 (4 months after the 4^th^ SPA).

### Serum Concentrations

At a given dose, peak and trough concentrations were highest in the IV groups and lowest in the SC groups. Peak geometric mean titres (GMT) after the first dose, as measured by ELISA, were noted on Day 3, with GMT (95% CI) of 28.98 μg/mL (25.16, 33.39) in the 2.5 mg/kg IV group, 58.51 μg/mL (49.41, 69.29) in the 5 mg/kg IV group, 257.22 μg/mL (127.52, 518.86) in the 20 mg/kg IV group, 10.84 μg/mL (8.84, 13.3) in the 2.5 mg/kg SC group, 22.78 μg/mL (20.05, 25.88) in the 5 mg/kg SC group, and 16.35 μg/mL (14.71, 18.18) in the 2.5 mg/kg IM group (**Figure 3; Supplemental Figure 1**). The lowest pre-administration troughs were noted on Day 112, just before the second infusion, with GMT (95% CI) of 3.4 μg/mL (2.51, 4.61) in the 2.5 mg/kg IV group, 6.5 μg/mL (5.6, 7.54) in the 5 mg/kg IV group, 27.2 μg/mL (23.85, 31.01) in the 20 mg/kg IV group, 0.97 μg/mL (0.65, 1.44) in the 2.5 mg/kg SC group, 3.09 μg/mL (2.2, 4.33) in the 5 mg/kg SC group, and 2.61 μg/mL (2.05, 3.31) in the 2.5 mg/kg IM group (Figure 3; **Supplemental Figure 1).**

**Figure 3.**
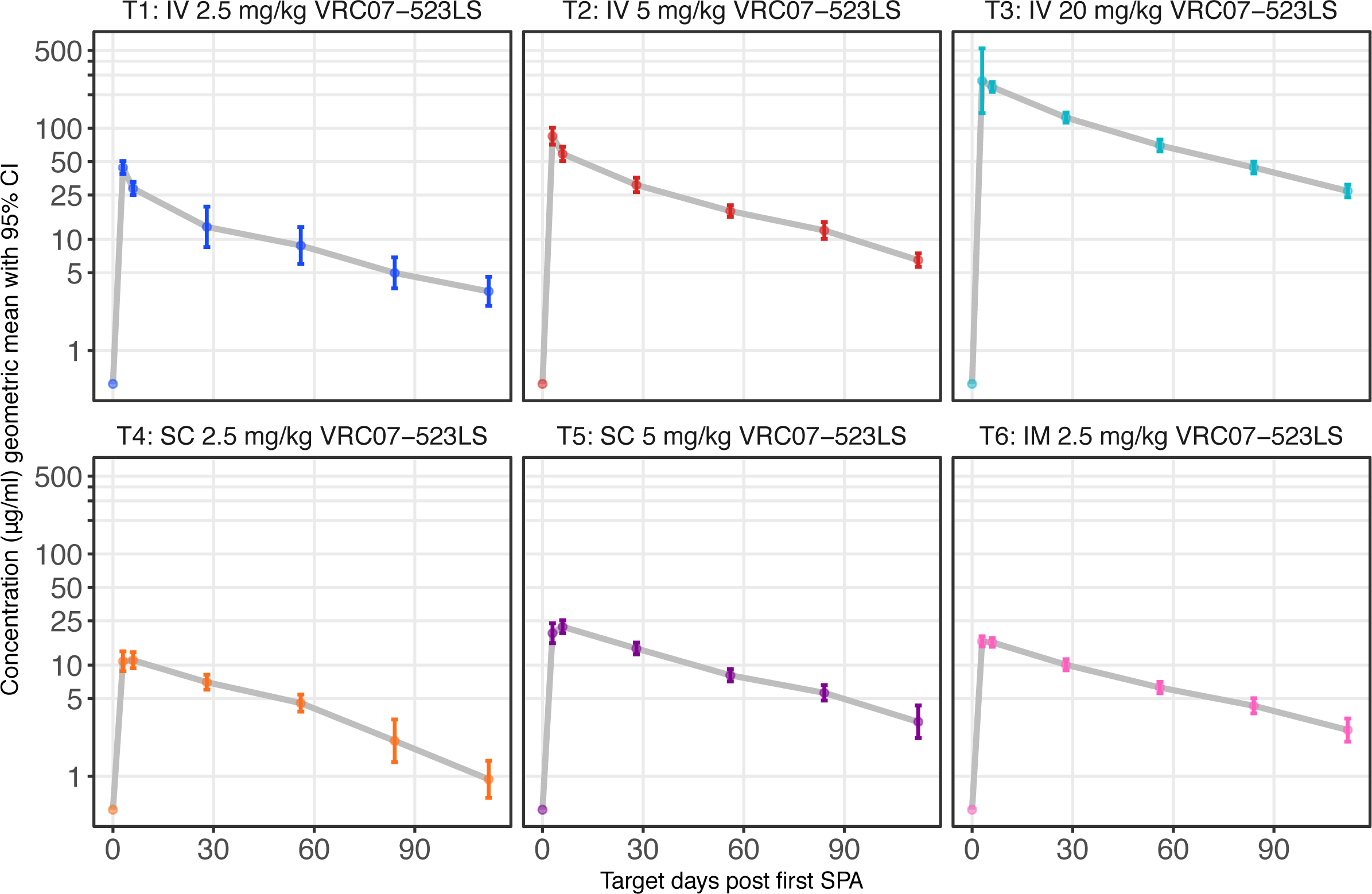
VRC07−523LS serum concentration (μg/mL) following first study product administration (SPA). Geometric means and 95% confidence intervals are presented, by treatment group and target day.

We compared the area under the curve (AUC) of the VRC07-523LS levels for the various doses and routes following the first administration. As a measure of exposure to VRC07-523LS, the AUC was higher in the 20 mg/kg IV group (**Supplemental Figure 2A**). When corrected for the administered dose, the three IV groups had higher AUCs than either the SC or the IM groups (**Supplemental Figure 2B**), reflecting the lower bioavailability of the SC and IM routes.

The subsequent four-month post-administration troughs were slightly higher than after the first dose and quite similar between the second and fifth study product administrations (**Figure 4; Supplemental Figure 3**). This result suggests that there was very little in vivo accumulation of VRC07-523LS with the 16-weekly administration schedule, at least up to five consecutive doses. Furthermore, the relatively consistent levels of VRC07-523LS measured between the second and fifth administrations given at a frequency of four months suggests that a near steady state is established after the second administration with peaks and troughs determined by the dose and route.

**Figure 4.**
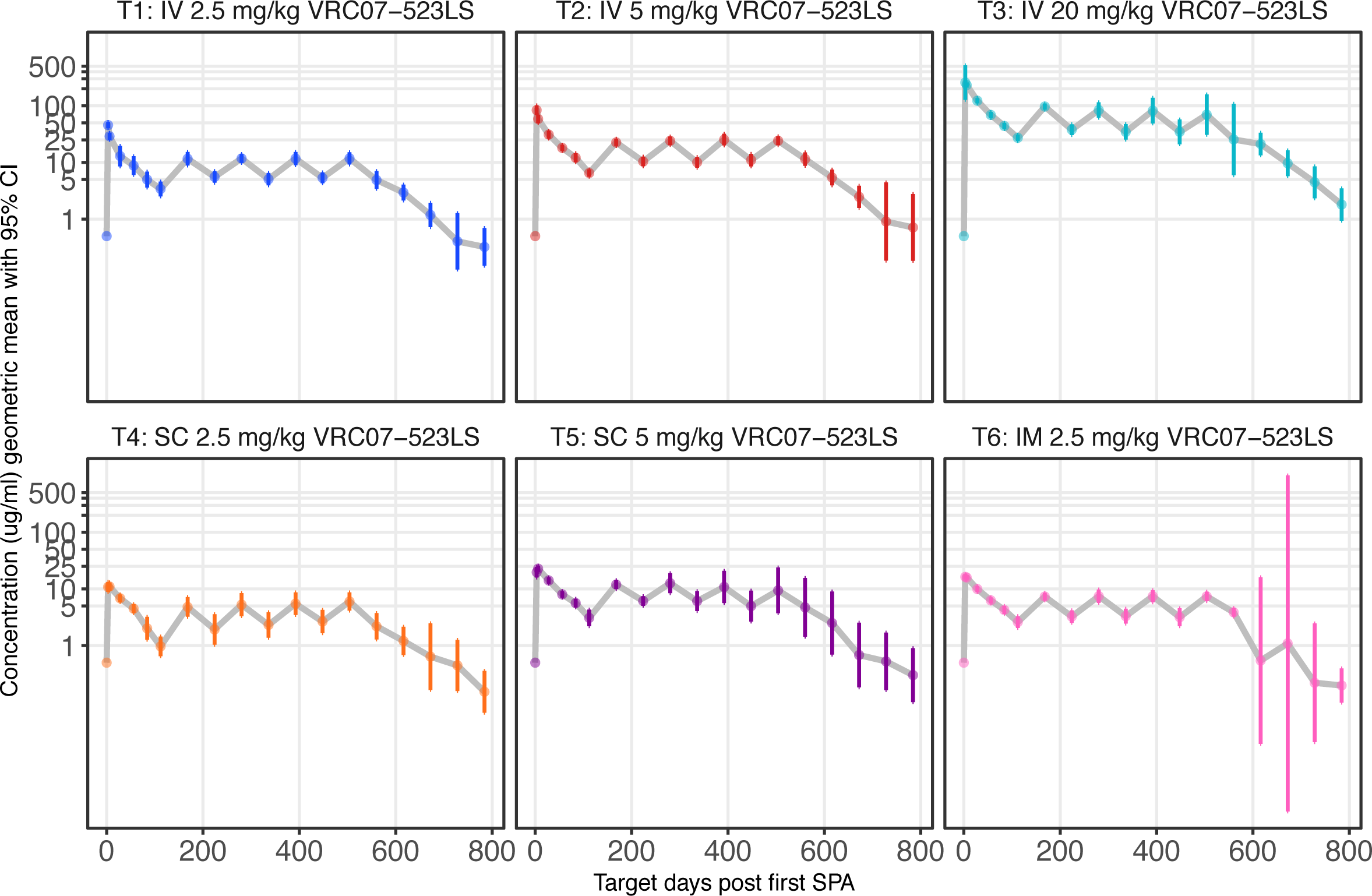
VRC07-523LS concentrations measured after product administration every four months at specified doses and routes. Peak levels were only assessed after the first dose. Levels following the second and subsequent doses were assessed by binding antigen multiplex assay (BAMA).

### Population PK Analysis

We conducted a population PK analysis of VRC07-523LS serum concentrations measured in healthy participants after up to five SPA via the IV, SC, or IM routes at various doses. Data were collected at 19 timepoints between Day 0 and Day 784, covering the period from baseline up to after the fifth SPA. The BICc determined that the two-compartment PK model was better at describing serum concentrations of VRC07-523LS than PK models with one or three compartments. The two-compartment PK model predicted well serum concentrations across routes of administration and doses (**Figure 5; Supplemental Figures 4A-F**). Estimated PK parameters are summarized in **Supplemental Table 3**.

**Figure 5.**
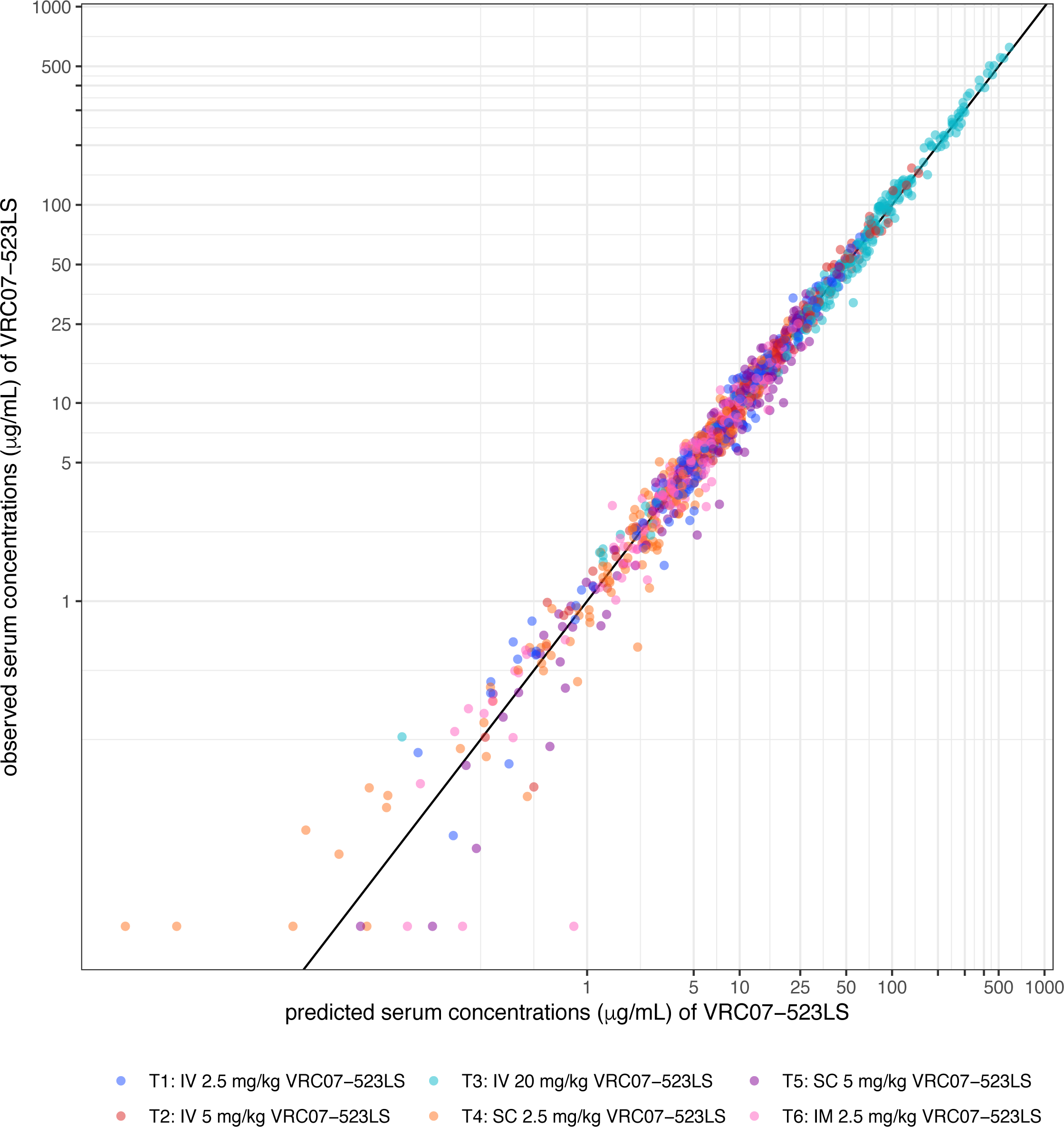
Measured serum concentrations of VRC07-523LS vs concentrations predicted by a two-compartment pharmacokinetic model.

Based on data measured between the first and second SPA using ELISA, the elimination half-life of VRC07-523LS was estimated as 42.43 days (SE = 0.75). Its bioavailability was estimated as 0.40 (SE = 0.0013) after SC administration, and 0.57 (SE = 0.03) after IM administration, as compared with IV administration. The difference between the bioavailability after SC and IM administrations was significant (p<0.001). The average clearance of VRC07-523LS was 0.11 L/d (SE = 0.0031).

The analyses of serum concentrations measured after the first SPA using ELISA versus after the second SPA using BAMA found significantly different PK parameters, namely estimated clearance (0.15 L/d versus 0.11 L/d; p < 0.001), central compartmental volume (3.18 versus 2.72, p < 0.001), peripheral compartmental volume (3.74 versus 3.59, p< 0.001), and inter-compartmental clearance (1.18 L/day versus 0.49 L/day; p < 0.001). The distribution and elimination half-lives were also significantly different (4.61 days versus 1.17 days, p < 0.001; and 42.67 days versus 42.43 days, p < 0.001) following the first SPA (by ELISA) and following the second SPA (by BAMA), respectively (**Supplemental Figure 5**). Although these differences are statistically significant, the absolute difference is quite small and unlikely to be clinically significant. Differences between the bioavailability and absorption rate constant based on BAMA versus ELISA derived data could not be estimated due to numerical instability occurring when fitting a population PK model that allowed for differences in these parameters between the two assays. This instability may be due to the differences in sampling frequency between the first and second SPA versus the less frequent sampling following the second SPA.

The analysis suggested that serum concentrations of VRC07-523LS increased linearly with the dose following IV and SC administrations, as evidenced by the population PK model properly fitting data across dose levels and routes of administration (**Figure 5; Supplemental Figures 4A-F**) and the similarity of the peak and trough levels as well as dose-normalized AUC across dose groups within a given route of administration (**Supplemental Figures 1 and 2**).

### Neutralization Activity

Neutralization activity was assayed at 3 time points (Weeks 8, 72, and 88 following the first infusion of study product) in a subset of approximately 10 participants in each active treatment group and 3 placebo recipients, as follows: 9 participants in Group 1, 10 in Group 2, 10 in Group 3, 10 in Group 4, 10 in Group 5, 11 in Group 6, and 3 in the Placebo group.

In the IV and SC groups, response magnitudes increased overall with dose at a given timepoint (**Figure 6; Supplemental Figure 6**). Overall, response rates and magnitudes at Week 72 (8 weeks after the fifth study product administration) were either comparable to or slightly higher than those measured at Week 8 (8 weeks after the first study product administration), again suggesting limited accumulation of VRC07-523LS between the first and fifth administrations. As expected, the lowest response rates and magnitudes were observed at Week 88 (24 weeks after the fifth study product administration).

**Figure 6.**
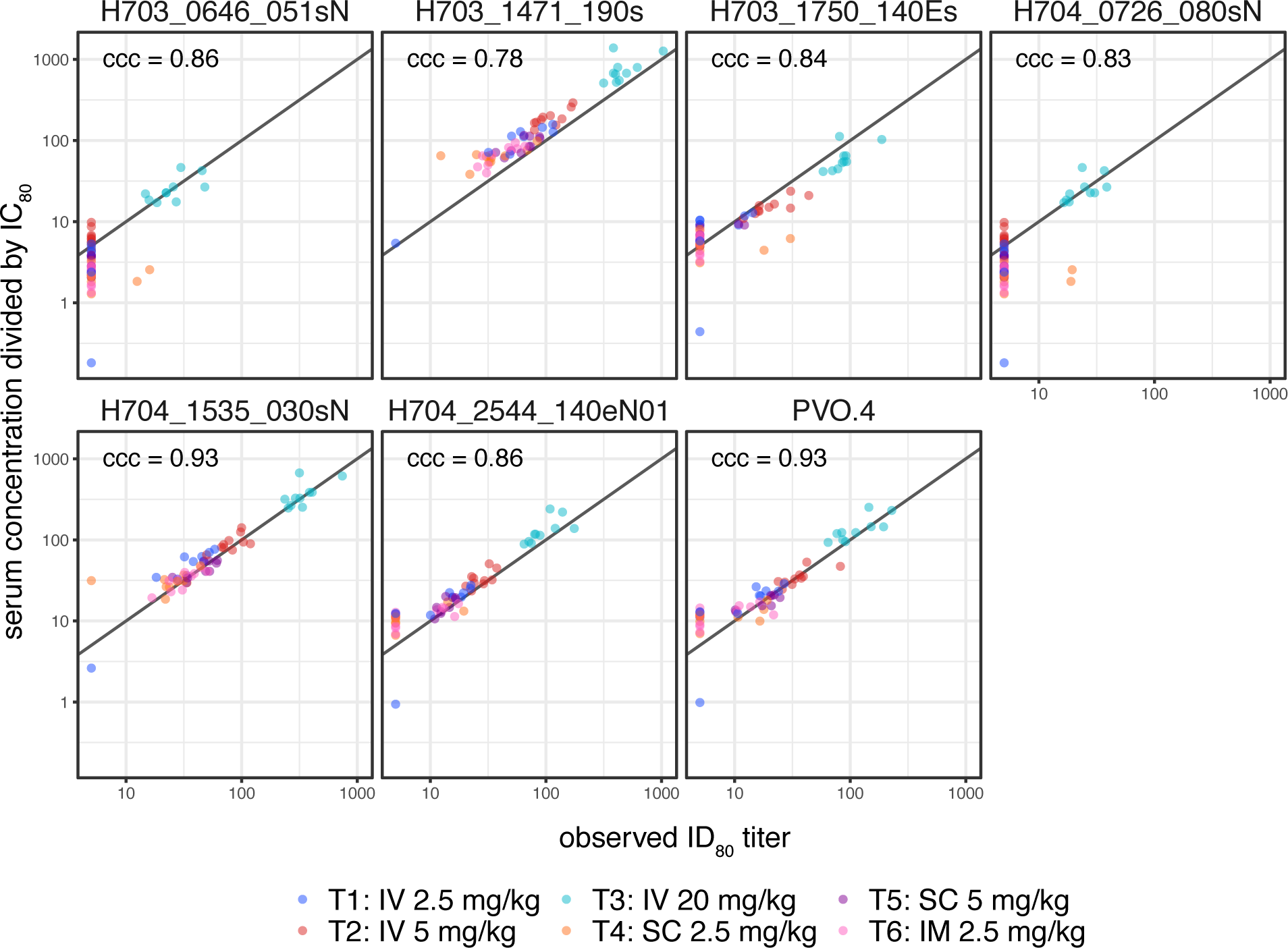
Neutralization activity of participant serum following VRC07-523LS administration against seven HIV-1 isolates collected from incident HIV-1 acquisition events of placebo recipients in the AMP trials. ID_80_ titre is shown.

In addition to confirming that neutralization activity of VRC07-523LS is preserved following in vivo administration, these results allow us to compare the established in vitro ID_80_ titre with the in vivo concentration to generate a predicted protective efficacy titre (PT_80_) (35). If an in vivo neutralization threshold of 200 is required for achieving potent prevention efficacy for VRC07-523LS (35), this level would only have been achieved for three of the primary HIV-1 isolates we tested even at 8 weeks after SPA (**Figure 6**).

At a given dose, higher serum neutralization titres were observed after IV administrations compared to either SC or IM administrations and this was particularly pronounced in the Magnitude-Breadth Area Under the Curve (MB-AUC) analysis (**Figure 7; Supplemental Figure 7**). The MB-AUC plots also indicate that neutralization activity was higher after receiving 5.0 mg/kg of VRC07-523LS via the SC route versus a lower dose of 2.5 mg/kg of VRC07-523LS via the IV route, suggesting that the levels of neutralization achieved by IV infusions may be reached by administering higher doses via the SC route. For the 2.5 mg/kg dose, which was the only one given via all three routes, the neutralization titres were highest for the IV route, followed by the IM route, and lowest via the SC route. As with the neutralization titres, response rates tended to be higher in the IV groups than in the SC or IM groups when comparing responses among groups that received the same dose.

**Figure 7.**
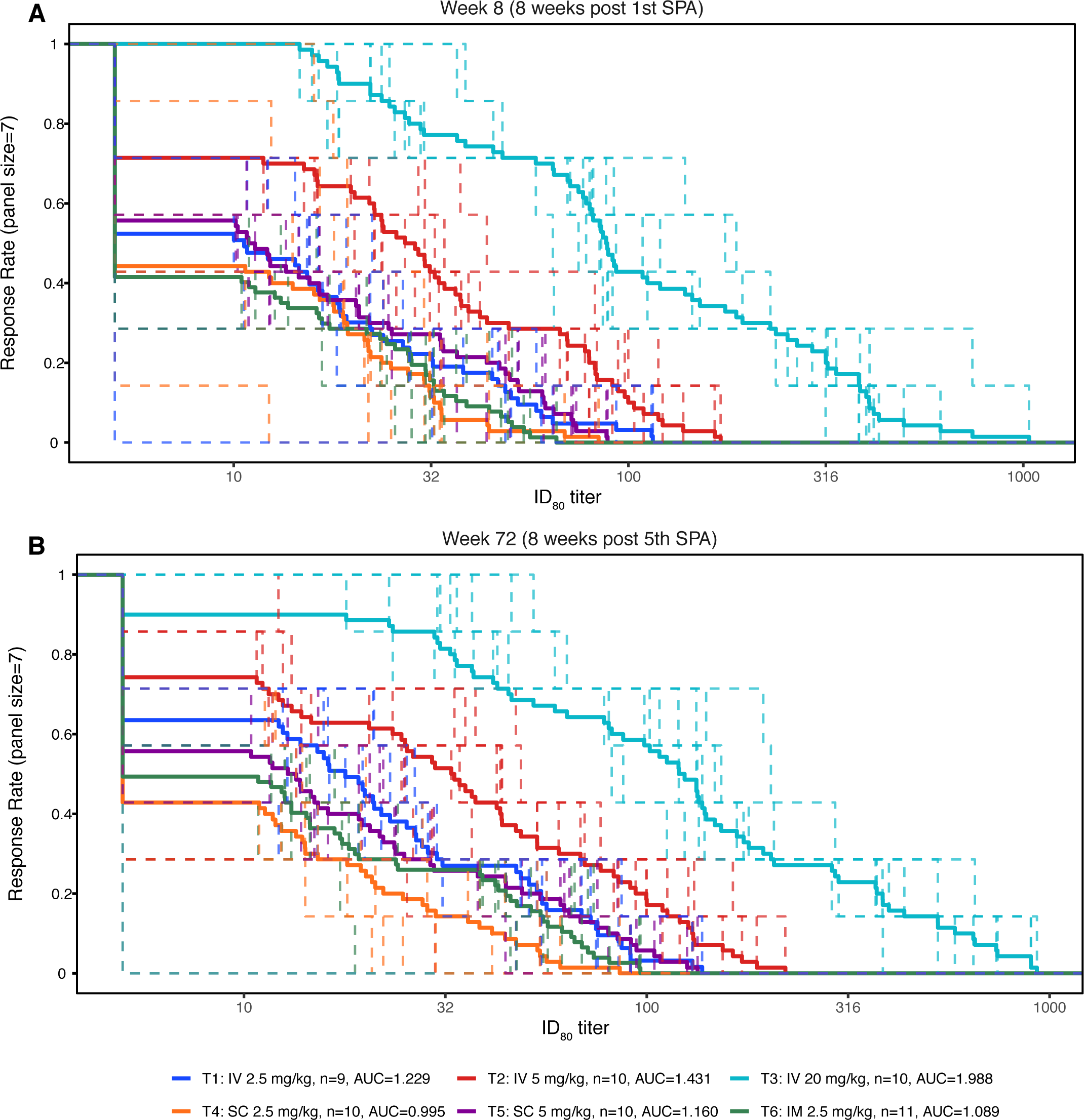
Magnitude-breadth curves for participant serum following VRC07-523LS administration against a panel of HIV-1 isolates collected from incident HIV-1 acquisition of placebo recipients in the AMP trials. ID_80_ titre is shown at Week 8 (A) and Week 72 (B).

## Discussion

This phase 1 clinical trial highlights several key findings which have implications for the design of future bnAb efficacy studies. First, a human neutralizing antibody engineered for breadth, potency, and a prolonged in vivo half-life was safe and generally well-tolerated when administered across a wide dose range and via different routes of administration. Second, although the half-life of this engineered bnAb is considerably longer than that of the VRC01 antibody used in the AMP studies (24), the half-life of VRC07-523LS is substantially less than VRC01LS (25). Third, while SC and IM administration of VRC07-523LS was safe and well-tolerated and these routes have notable operational advantages (11), decreased bioavailability may have implications for achieving target concentrations, and inter-individual variability in PK parameters observed must be considered in designing a combination bnAb efficacy study with VRC07-523LS.

In this study, we found that the serum concentrations of VRC07-523LS increased linearly across the dose range administered in this study. We also found that the peak and trough concentrations were highest in the IV groups and lowest in the SC groups at any given dose. There was also no significant difference in the half-life of VRC07-523LS when administered by different routes. These trends were also true for VRC01 and VRC01LS in other studies (25, 26), as well as for other bnAbs (27), which likely reflects generalizable PK behavior of antiviral antibodies in unexposed hosts. Some studies noted shorter half-lives of anti-HIV-1 bnAbs in persons living with HIV (28–30), which has implications for use of these biologics in HIV treatment and cure studies.

We also found an excellent correlation between the predicted level of VRC07-523LS and the measured serum concentrations, suggesting that a two-compartment model can predict the PK of this monoclonal antibody following repeated IV, SC, or IM administrations. This correlation was robust to dose, delivery route, and method of antibody measurement (ELISA vs BAMA). The high performance of these prediction models may allow future phase 1 studies to be smaller and more efficient, in terms of the number of participants needed, the number of injections or infusions administered, and the total length of follow-up needed to develop a robust PK model. This finding in turn has the potential to facilitate moving bnAbs into efficacy studies.

Data from this study has informed ongoing studies of combination bnAbs as it is likely that combinations of bnAbs targeting complementary epitopes on Env will be necessary to provide sufficient protection in vivo (6). VRC07-523LS has been evaluated in combination with the non-LS-mutated bnAbs PGT121, PGDM1400, and 10-1074, in the HVTN 130/HPTN 089 study (31). PGT121 and 10-1074 target the V3 glycan epitope whereas PGDM1400 targets the V1V2 region, and all three, while narrower in breadth than VRC07-523LS, are considerably more potent (32). As bnAbs with longer half-lives are likely to be more feasible for deployment (5, 6), VRC07-523LS is also being evaluated in combination with an engineered version of PGT121 called PGT121.414.LS in the recently completed HVTN 136/HPTN 092 trial [NCT04212091]. In the HVTN 140/HPTN 101 study [NCT05184452], VRC07-523LS and PGT121.414.LS were tested in combination with PGDM1400LS. In the multi-phase CAPRISA 012 studies (33, 34), VRC07-523LS was tested with CAP256V2LS, a V1V2 targeting bnAb which is particularly potent against clade C isolates of HIV-1. Some of these studies include modified SC administration methods (for example, SC infusion via pump in HVTN 136/HPTN 092 [NCT04212091] and co-administration of bnAbs with hyaluronidase in CAPRISA 012 (33, 34)) to try to increase the bioavailability of the SC route. Furthermore, analysis of mucosal tissue and secretion PK of VRC07-523LS at rectal and vaginal sites is ongoing in the HVTN 128 trial [NCT03735849], which is particularly relevant as the vast majority of HIV-1 infections occur at mucosal sites.

Recently, the correlates of risk analyses from the AMP studies were used to generate a model of protective efficacy (PT_80_) that can be used for predicting efficacy of bnAbs against HIV-1 (35). The PT_80_ model combines in vitro IC_80_ neutralization data with in vivo PK data to provide an estimate of protection against a population of viruses to which participants may be exposed (35). This model was used to predict prevention efficacy (PE) of a hypothetical repeat of the AMP studies using the broader and more potent mAb VRC07-523LS as a single agent and suggested that PE in this case could have been approximately 79% against clade C viruses (35). However, the PT_80_ model also suggested that a high-dose three bnAb combination regimen including VRC07-523LS could have a PE of approximately 95% against a panel of recent clade C isolates (35). Ultimately, large efficacy studies will need to be performed with combination bnAbs to assess their ability to prevent HIV-1 acquisition.

UNAIDS recently released their preferred product characteristics (PPC) for mAbs for HIV-1 prevention, noting that longer-acting mAbs could play an important role in future pre-exposure prophylaxis (PrEP) campaigns as many people may opt for an injectable product rather than anti-retroviral pills or intra-vaginal rings (11). Long-acting prevention methods may be desirable for key populations such as pregnant women and adolescent girls who are at high risk of HIV-1 acquisition, particularly in sub-Saharan Africa (11). While the UNAIDS PPC expressed a preference for SC administration, as noted for other low-volume mAbs for non-infectious indications (36), the participants in our SC groups had a somewhat less enthusiastic opinion about acceptability of SC administration compared to the other groups. Furthermore, the lower bioavailability observed for VRC07-523LS combined with the high concentrations likely needed to exceed the PT_80_ for circulating strains of HIV-1 argue that IV administration is more likely to be successful for HIV-1 prevention until the technical hurdles which impact injection volume and potency are overcome.

## Data Availability

Upon acceptance, the data underlying the findings of this manuscript will be made publicly available at the public-facing HVTN website (https://atlas.scharp.org/).

## Acknowledgments

We wish to thank our volunteers for their generous participation in this study as well as the dedicated staff at the HVTN and HPTN clinical research sites and affiliated laboratories who made the study possible. We also wish to thank Mindy Miner and Nicole Na for editorial assistance as well as Jessica Fogel and Fatima Laher for their thoughtful comments on the manuscript.

The HVTN 127/HPTN 087 Study Group: In addition to the authors of this article, members of the HVTN 127/HPTN 087 Study Group consist of: Myron Cohen, Lawrence Corey, Jill Zeller, Gina Escamilla, Gail Broder, April Randhawa, Jessica Andriesen, Amanda Brown, Carter Bentley, Jontraye Davis, Huiyin Lu, Jignesh Patel, Tahereh Nourbakhsh, Shannon Grant, Rotrease Reagan, Jamel Young, Noshima Darden-Tabb, Jorge Benitez, Brett Gray, Joshua A. Weiner, Gabriela Kovacikova, Chanc E, VanWinkle Orzell, Maureen Furlong, Nathan Erdmann, Sonya Heath, Michel Obeid, Rosemary Hottinger, Rossi Fish, Adrianna Boulin, Gary Daffin, Marcy Gelman, Colleen Kelley, Jane A. Kleinjan, Jon A. Gothing, Elvin J. Fontana-Martinez, Michael S. Seaman, Raphael Dolin, and Jennifer A. Johnson.

## Notes

### Competing Interest Statement

SRW has conducted clinical trials funded by Janssen Vaccines, Moderna, Pfizer, Vir, Worcester HIV Vaccine, and Sanofi Pasteur, and serves on Independent Data Monitoring Committees for Janssen Vaccines and BioNTech SRW?s spouse is an employee of Regeneron Pharmaceuticals and may hold stock and/or stock options. CLG has received research support from Gilead and ViiV Healthcare and conducted clinical trials funded by Moderna and Novavax. MEA has received research support from Janssen, Be Bio, and Moderna.

### Clinical Trial

NCT03387150

### Funding Statement

US National Institutes of Health (NIH) [www.nih.gov] grants UM1 AI068614 (STK, CK, CAP, SJ, GDT), UM1 AI068635 (OH, ZH, HL, YH), UM1 AI068618 (KS, GDT, DCM), UM1 AI069412 (SRW, LRB), UL1 RR025758 (LRB), P30 AI50410 (CLG), UM1 AI069423 (CLG), UM1 AI068619 (PA, JD), UM1 AI068617 (TS), UM1 AI068613 (EPM), UM1 TR004406 (CLG). JAGB, LLP, and WC are/were Medical Officers with the National Institute of Allergy and Infectious Diseases (NIAID)/NIH and ABM, BF, LG, and JCM are/were Staff Scientists with NIAID/NIH. The funder had no other role in the study design, data collection and analysis, decision to publish, or preparation of the manuscript.

### Author Declarations

The Partners Human Research Committee of Brigham and Women's Hospital gave ethical approval of this work. The Centre Hospitalier Universitaire Vaudois Institutional Review Board of the Centre Hospitalier Universitaire Vaudois gave ethical approval of this work. The Fred Hutchinson Cancer Research Center Institutional Review Board gave ethical approval of this work done at the University of North Carolina at Chapel Hill, the Ponce de Leon/Emory University, and the University of Alabama at Birmingham. The Fenway Health Institutional Review Board of the Fenway Institute gave ethical approval of this work.

