## Supplemental Material for "A Randomised Clinical Trial of the Safety and Pharmacokinetics of VRC07-523LS Administered via Different Routes and Doses (HVTN 127/HPTN 087)"

| | Isolate | VRC07-523LS | VRC01 |
|---|---|---|---|
| $IC_{50}$ (µg/mL) | H703_0646_051sN | 0.98 | 7.38 |
| | H703_1471_190s | 0.03 | 0.17 |
| | H703_1750_140Es | 0.32 | 0.99 |
| | H704_0726_080sN | 0.91 | 4.7 |
| | H704_1535_030sN | 0.06 | 0.1 |
| | H704_2544_140eN01 | 0.16 | 0.79 |
| | PVO.4 | 0.17 | 0.54 |
| $IC_{80}$ (µg/mL) | H703_0646_051sN | 2.75 | 20.45 |
| | H703_1471_190s | 0.09 | 0.41 |
| | H703_1750_140Es | 1.14 | 2.66 |
| | H704_0726_080sN | 2.76 | 15.5 |
| | H704_1535_030sN | 0.19 | 0.27 |
| | H704_2544_140eN01 | 0.53 | 2.48 |
| | PVO.4 | 0.51 | 1.32 |

**Supplemental Table 1.** In vitro $IC_{50}$ and $IC_{80}$ of the clinical lot of two bnAbs (VRC07-523LS and VRC01) against seven Env-pseudotyped viruses. The *env* sequences are derived from isolates sequenced from incident acquisition events from the AMP studies.

### Supplemental Table 2A: Discomfort acceptability of IV, SC, and IM infusion

| Question | Route | Visit | Visit description | No discomfort | Discomfort acceptable | Discomfort not acceptable |
|---|---|---|---|---|---|---|
| Level of discomfort from [IV infusion/SC injection(s)/IM injection(s)] | IV | Day 0 | 1st SPA | 52/59 88.1% | 6/59 10.2% | 1/59 1.7% |
| | | Month 4 | 2nd SPA | 46/55 83.6% | 9/55 16.4% | 0/55 0.0% |
| | | Month 8 | 3rd SPA | 43/51 84.3% | 8/51 15.7% | 0/51 0.0% |
| | | Month 12 | 4th SPA | 41/49 83.7% | 8/49 16.3% | 0/49 0.0% |
| | | Month 16 | 5th SPA | 38/44 86.4% | 6/44 13.6% | 0/44 0.0% |
| | SC | Day 0 | 1st SPA | 22/41 53.7% | 18/41 43.9% | 1/41 2.4% |
| | | Month 4 | 2nd SPA | 13/40 32.5% | 26/40 65.0% | 1/40 2.5% |
| | | Month 8 | 3rd SPA | 19/37 51.4% | 18/37 48.6% | 0/37 0.0% |
| | | Month 12 | 4th SPA | 14/33 42.4% | 18/33 54.5% | 1/33 3.0% |
| | | Month 16 | 5th SPA | 15/31 48.4% | 15/31 48.4% | 1/31 3.2% |
| | IM | Day 0 | 1st SPA | 16/24 66.7% | 8/24 33.3% | 0/24 0.0% |
| | | Month 4 | 2nd SPA | 12/23 52.2% | 11/23 47.8% | 0/23 0.0% |
| | | Month 8 | 3rd SPA | 15/23 65.2% | 8/23 34.8% | 0/23 0.0% |
| | | Month 12 | 4th SPA | 14/20 70.0% | 6/20 30.0% | 0/20 0.0% |
| | | Month 16 | 5th SPA | 14/20 70.0% | 6/20 30.0% | 0/20 0.0% |

### Supplemental Table 2B: Pain acceptability of IV, SC, and IM infusion

| Question | Route | Visit | Visit description | No pain | Pain acceptable | Pain not acceptable |
|---|---|---|---|---|---|---|
| Level of pain from [IV infusion/SC injection(s)/IM injection(s)] | IV | Day 0 | 1st SPA | 54/59 91.5% | 5/59 8.5% | 0/59 0.0% |
| | | Month 4 | 2nd SPA | 50/55 90.9% | 4/55 7.3% | 1/55 1.8% |
| | | Month 8 | 3rd SPA | 46/51 90.2% | 5/51 9.8% | 0/51 0.0% |
| | | Month 12 | 4th SPA | 45/49 91.8% | 4/49 8.2% | 0/49 0.0% |
| | | Month 16 | 5th SPA | 40/44 90.9% | 4/44 9.1% | 0/44 0.0% |
| | SC | Day 0 | 1st SPA | 20/41 48.8% | 20/41 48.8% | 1/41 2.4% |
| | | Month 4 | 2nd SPA | 21/40 52.5% | 18/40 45.0% | 1/40 2.5% |
| | | Month 8 | 3rd SPA | 20/37 54.1% | 16/37 43.2% | 1/37 2.7% |
| | | Month 12 | 4th SPA | 14/33 42.4% | 17/33 51.5% | 2/33 6.1% |
| | | Month 16 | 5th SPA | 17/31 54.8% | 13/31 41.9% | 1/31 3.2% |
| | IM | Day 0 | 1st SPA | 12/24 50.0% | 12/24 50.0% | 0/24 0.0% |
| | | Month 4 | 2nd SPA | 12/23 52.2% | 10/23 43.5% | 1/23 4.3% |
| | | Month 8 | 3rd SPA | 12/23 52.2% | 11/23 47.8% | 0/23 0.0% |
| | | Month 12 | 4th SPA | 11/20 55.0% | 9/20 45.0% | 0/20 0.0% |
| | | Month 16 | 5th SPA | 14/20 70.0% | 6/20 30.0% | 0/20 0.0% |

### Supplemental Table 2C: Anxiety acceptability of IV, SC, and IM infusion

| Question | Route | Visit | Visit description | No anxiety | Anxiety acceptable | Anxiety not acceptable |
|---|---|---|---|---|---|---|
| Level of anxiety from [IV infusion/SC injection(s)/IM injection(s)] | IV | Day 0 | 1st SPA | 56/59 94.9% | 3/59 5.1% | 0/59 0.0% |
| | | Month 4 | 2nd SPA | 51/55 92.7% | 4/55 7.3% | 0/55 0.0% |
| | | Month 8 | 3rd SPA | 46/51 90.2% | 5/51 9.8% | 0/51 0.0% |
| | | Month 12 | 4th SPA | 45/49 91.8% | 4/49 8.2% | 0/49 0.0% |
| | | Month 16 | 5th SPA | 43/44 97.7% | 1/44 2.3% | 0/44 0.0% |
| | SC | Day 0 | 1st SPA | 30/41 73.2% | 11/41 26.8% | 0/41 0.0% |
| | | Month 4 | 2nd SPA | 35/40 87.5% | 5/40 12.5% | 0/40 0.0% |
| | | Month 8 | 3rd SPA | 35/37 94.6% | 2/37 5.4% | 0/37 0.0% |
| | | Month 12 | 4th SPA | 29/33 87.9% | 3/33 9.1% | 1/33 3.0% |
| | | Month 16 | 5th SPA | 27/31 87.1% | 4/31 12.9% | 0/31 0.0% |
| | IM | Day 0 | 1st SPA | 19/24 79.2% | 5/24 20.8% | 0/24 0.0% |
| | | Month 4 | 2nd SPA | 20/23 87.0% | 3/23 13.0% | 0/23 0.0% |
| | | Month 8 | 3rd SPA | 23/23 100.0% | 0/23 0.0% | 0/23 0.0% |
| | | Month 12 | 4th SPA | 18/20 90.0% | 2/20 10.0% | 0/20 0.0% |
| | | Month 16 | 5th SPA | 17/20 85.0% | 3/20 15.0% | 0/20 0.0% |

Supplemental Table 2D: Amount of time acceptability of IV, SC, and IM infusion

| Question | Route | Visit | Visit description | Amount of time acceptable | Amount of time not acceptable |
|---|---|---|---|---|---|
| The amount of time required to spend in the clinic during the [IV infusion/SC injection(s)/IM injection(s)] | IV | Day 0 | 1st SPA | 54/59 91.5% | 5/59 8.5% |
| | | Month 4 | 2nd SPA | 54/55 98.2% | 1/55 1.8% |
| | | Month 8 | 3rd SPA | 48/51 94.1% | 3/51 5.9% |
| | | Month 12 | 4th SPA | 47/49 95.9% | 2/49 4.1% |
| | | Month 16 | 5th SPA | 43/44 97.7% | 1/44 2.3% |
| | SC | Day 0 | 1st SPA | 41/41 100.0% | 0/41 0.0% |
| | | Month 4 | 2nd SPA | 39/40 97.5% | 1/40 2.5% |
| | | Month 8 | 3rd SPA | 35/37 94.6% | 2/37 5.4% |
| | | Month 12 | 4th SPA | 29/33 87.9% | 4/33 12.1% |
| | | Month 16 | 5th SPA | 27/31 87.1% | 4/31 12.9% |
| | IM | Day 0 | 1st SPA | 22/24 91.7% | 2/24 8.3% |
| | | Month 4 | 2nd SPA | 23/23 100.0% | 0/23 0.0% |
| | | Month 8 | 3rd SPA | 23/23 100.0% | 0/23 0.0% |
| | | Month 12 | 4th SPA | 20/20 100.0% | 0/20 0.0% |
| | | Month 16 | 5th SPA | 20/20 100.0% | 0/20 0.0% |

Supplemental Table 2E: Willingness to use of IV, SC, and IM infusion in general population

| Question | Route | Visit | Visit description | Very willing | Somewhat willing | Not willing |
|---|---|---|---|---|---|---|
| How willing would you be to get [IV infusions/SC injections/IM injection(s)] to prevent you from getting a serious disease, such as HIV, if you were at risk for that disease? | IV | Day 0 | 1st SPA | 47/59 79.7% | 12/59 20.3% | 0/59 0.0% |
| | | Month 4 | 2nd SPA | 46/55 83.6% | 9/55 16.4% | 0/55 0.0% |
| | | Month 8 | 3rd SPA | 41/51 80.4% | 10/51 19.6% | 0/51 0.0% |
| | | Month 12 | 4th SPA | 41/49 83.7% | 8/49 16.3% | 0/49 0.0% |
| | | Month 16 | 5th SPA | 37/44 84.1% | 7/44 15.9% | 0/44 0.0% |
| | SC | Day 0 | 1st SPA | 35/41 85.4% | 5/41 12.2% | 1/41 2.4% |
| | | Month 4 | 2nd SPA | 32/40 80.0% | 8/40 20.0% | 0/40 0.0% |
| | | Month 8 | 3rd SPA | 32/37 86.5% | 5/37 13.5% | 0/37 0.0% |
| | | Month 12 | 4th SPA | 27/33 81.8% | 6/33 18.2% | 0/33 0.0% |
| | | Month 16 | 5th SPA | 24/31 77.4% | 6/31 19.4% | 1/31 3.2% |
| | IM | Day 0 | 1st SPA | 22/24 91.7% | 2/24 8.3% | 0/24 0.0% |
| | | Month 4 | 2nd SPA | 22/23 95.7% | 1/23 4.3% | 0/23 0.0% |
| | | Month 8 | 3rd SPA | 22/23 95.7% | 1/23 4.3% | 0/23 0.0% |
| | | Month 12 | 4th SPA | 20/20 100.0% | 0/20 0.0% | 0/20 0.0% |
| | | Month 16 | 5th SPA | 20/20 100.0% | 0/20 0.0% | 0/20 0.0% |

Supplemental Table 2F: Recommendations for use of IV, SC, and IM infusion in general population

| Question | Route | Visit | Visit description | Yes | No | Don't know |
|---|---|---|---|---|---|---|
| Would you recommend receiving the [IV infusion/SC injection(s)/IM injection(s)] to a friend who is at risk for HIV? | IV | Day 0 | 1st SPA | 54/59 91.5% | 1/59 1.7% | 4/59 6.8% |
| | | Month 4 | 2nd SPA | 52/55 94.5% | 0/55 0.0% | 3/55 5.5% |
| | | Month 8 | 3rd SPA | 49/51 96.1% | 0/51 0.0% | 2/51 3.9% |
| | | Month 12 | 4th SPA | 47/49 95.9% | 1/49 2.0% | 1/49 2.0% |
| | | Month 16 | 5th SPA | 43/44 97.7% | 0/44 0.0% | 1/44 2.3% |
| | SC | Day 0 | 1st SPA | 40/41 97.6% | 1/41 2.4% | 0/41 0.0% |
| | | Month 4 | 2nd SPA | 38/40 95.0% | 1/40 2.5% | 1/40 2.5% |
| | | Month 8 | 3rd SPA | 34/37 91.9% | 0/37 0.0% | 3/37 8.1% |
| | | Month 12 | 4th SPA | 30/33 90.9% | 1/33 3.0% | 2/33 6.1% |
| | | Month 16 | 5th SPA | 24/31 77.4% | 3/31 9.7% | 4/31 12.9% |
| | IM | Day 0 | 1st SPA | 24/24 100.0% | 0/24 0.0% | 0/24 0.0% |
| | | Month 4 | 2nd SPA | 23/23 100.0% | 0/23 0.0% | 0/23 0.0% |
| | | Month 8 | 3rd SPA | 23/23 100.0% | 0/23 0.0% | 0/23 0.0% |
| | | Month 12 | 4th SPA | 20/20 100.0% | 0/20 0.0% | 0/20 0.0% |
| | | Month 16 | 5th SPA | 19/20 95.0% | 0/20 0.0% | 1/20 5.0% |

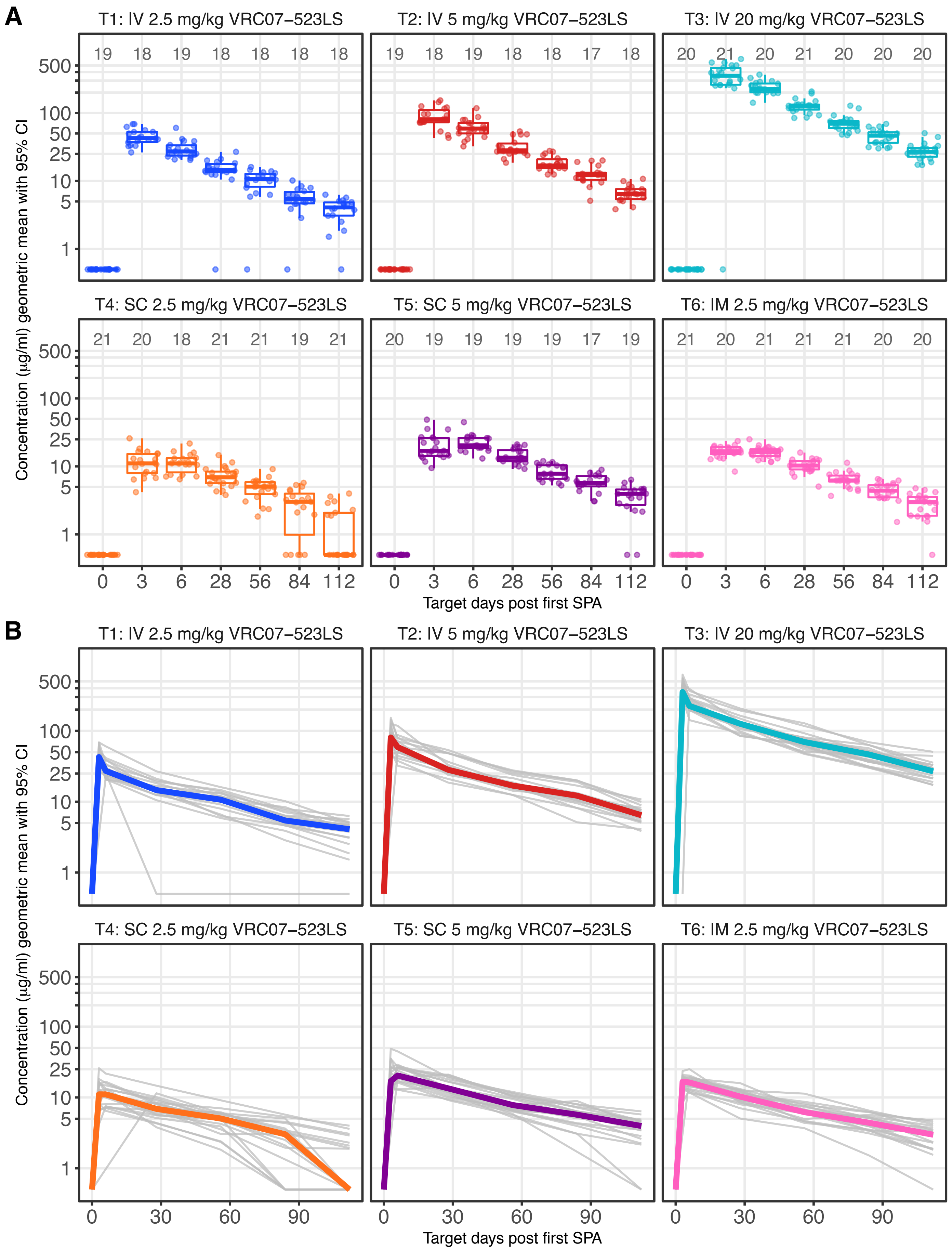

**Supplemental Figure 1**. VRC07-523LS levels following first study product administration (SPA). Individual measurements are shown in Panel **A**; note X-axis is not to scale. Longitudinal per-participant levels are shown in Panel **B**.

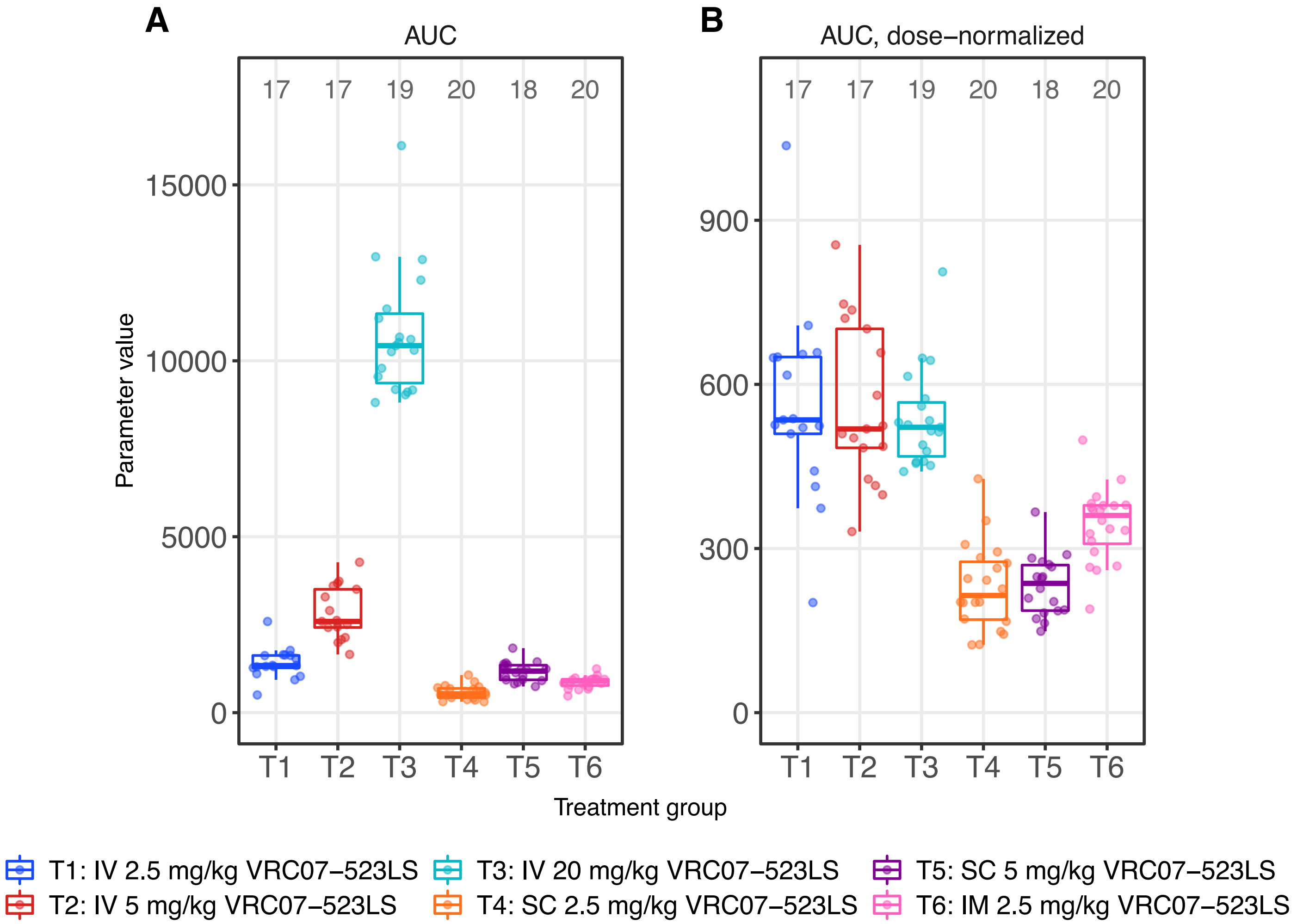

**Supplemental Figure 2.** VRC07-523LS serum concentrations were measured following the first study product administration and the area under the curve (AUC, panel **A**) and AUC corrected for dose (**B**) were computed to characterize the pharmacokinetics of VRC07-523LS.

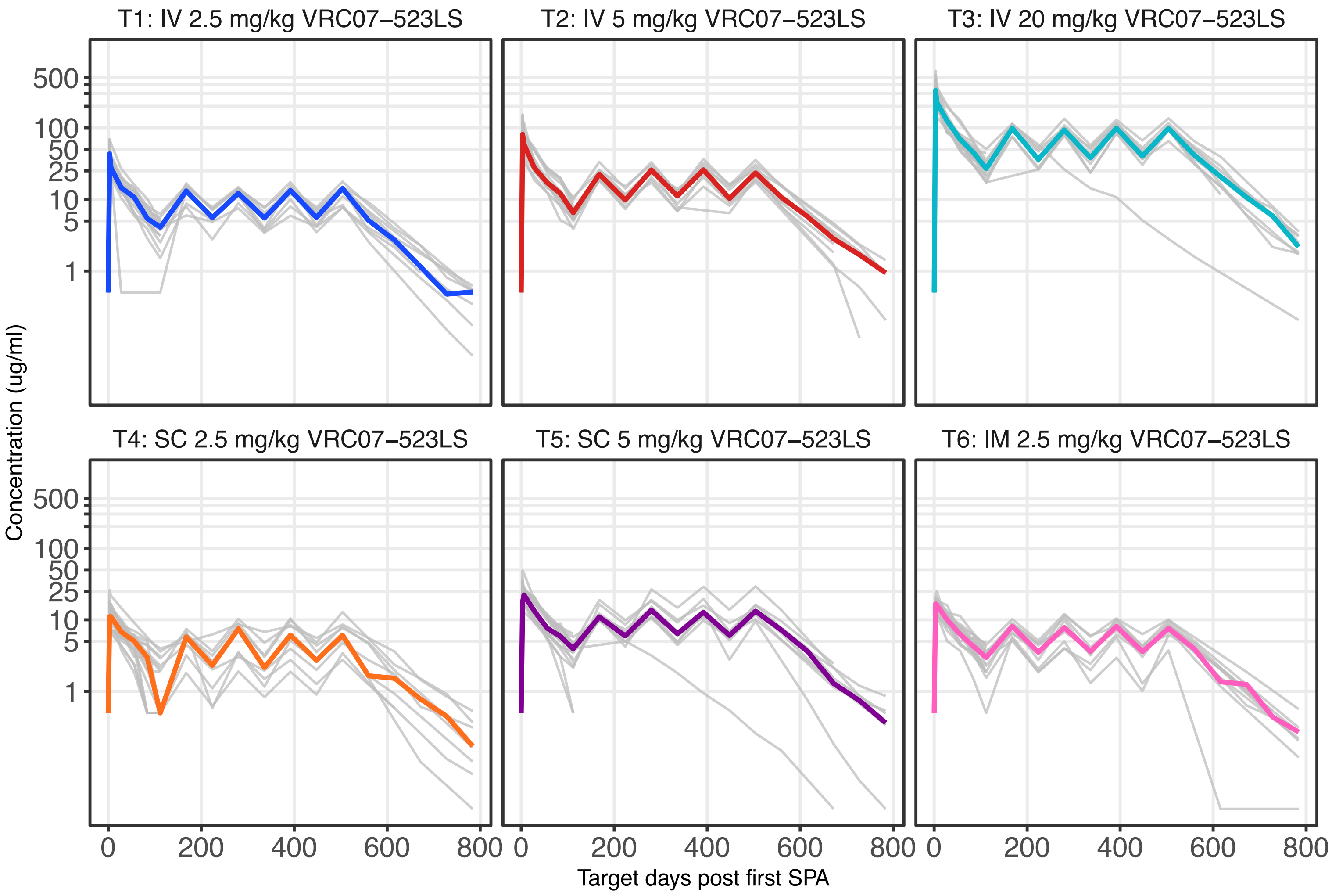

**Supplemental Figure 3.** VRC07-523LS concentrations measured after product administration every four months at specified doses and routes. Peak levels were only assessed after the first dose. Levels following the the second and subsequent doses were assessed by binding antigen multiplex assay (BAMA). Individual-level data are shown in grey and the group median is shown in colour.

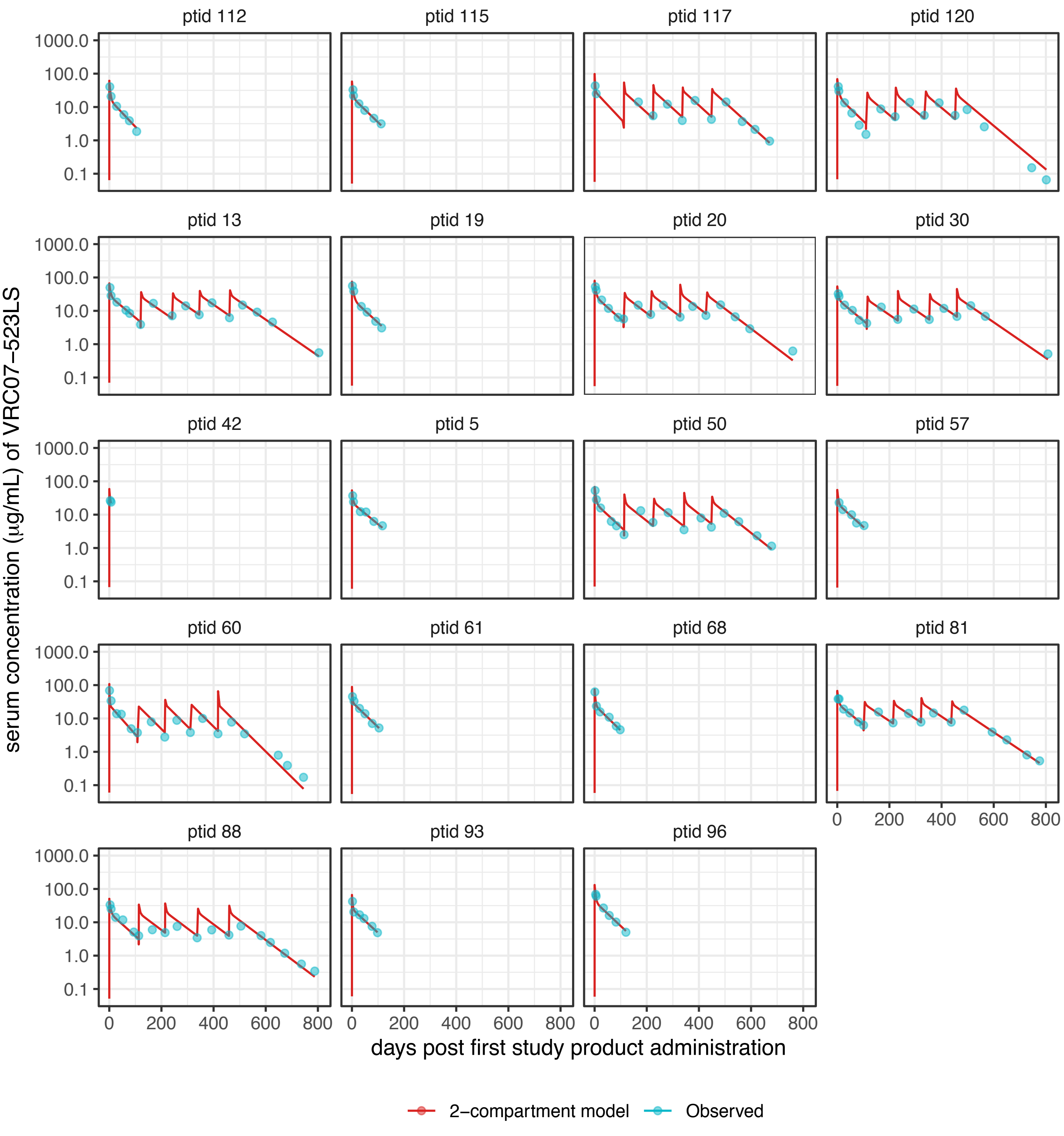

**Supplemental Figure 4A**. Observed (·) and predicted (–) serum concentrations of VRC07–523LS as a function of time in individual participants (one per plot); Group 1 (IV 2.5 mg/kg) is shown. The two–compartment population PK model with fully unstructured random effects variance–covariance matrix was fitted to VRC07–523LS concentrations.

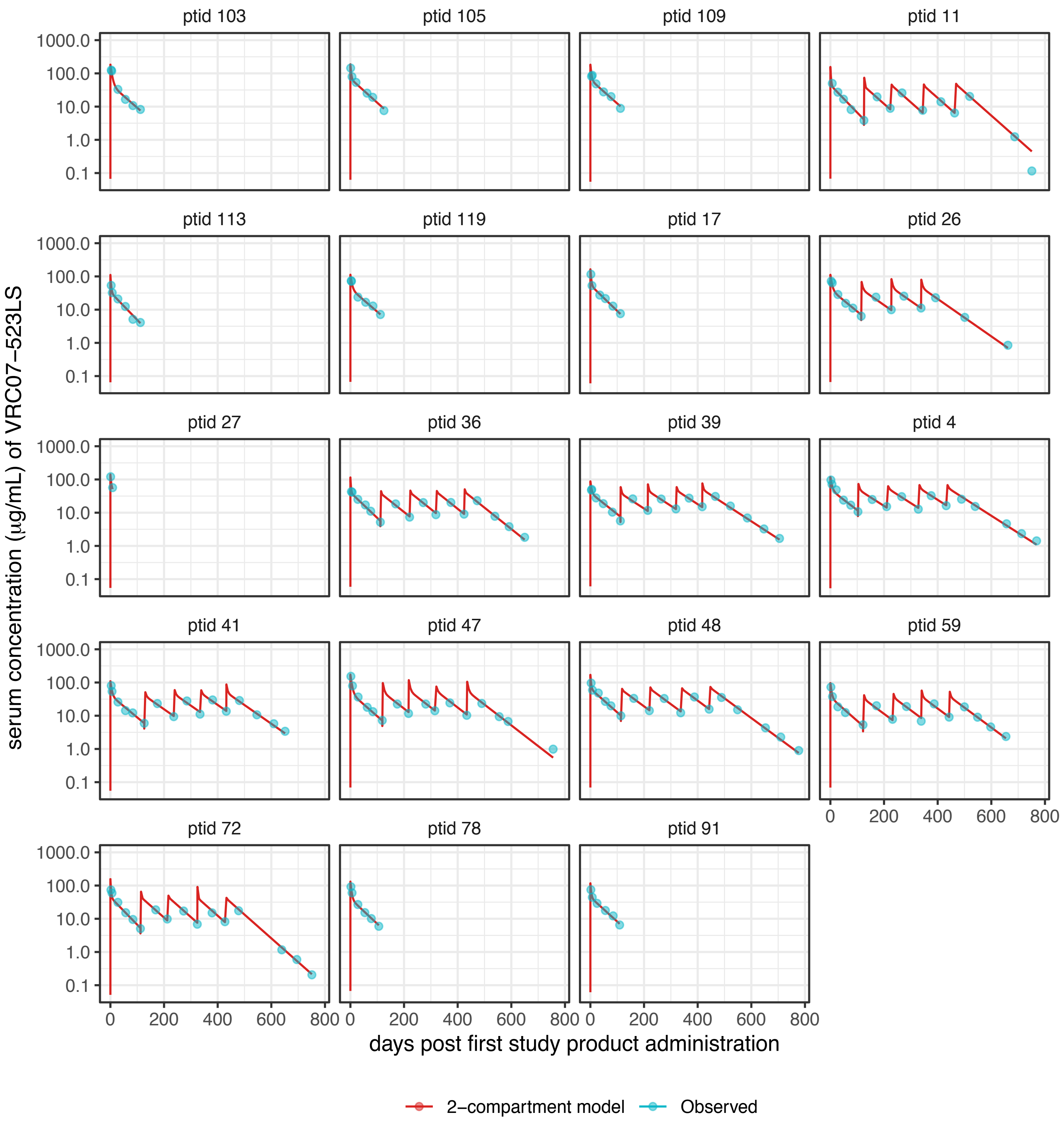

**Supplemental Figure 4B**. Observed (·) and predicted (–) serum concentrations of VRC07−523LS as a function of time in individual participants (one per plot); Group 2 (IV 5 mg/kg) is shown. The two−compartment population PK model with fully unstructured random effects variance−covariance matrix was fitted to VRC07−523LS concentrations.

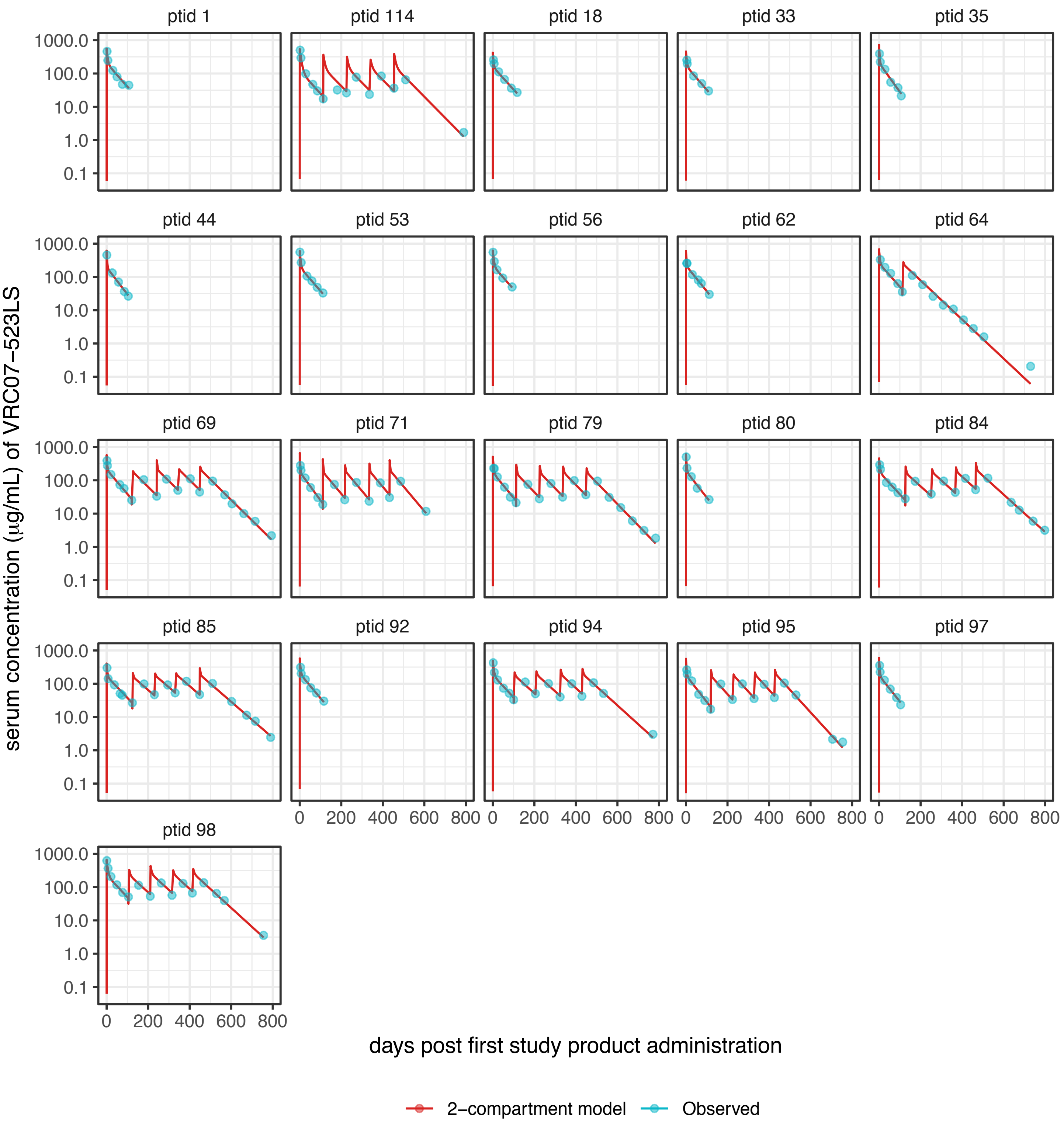

**Supplemental Figure 4C**. Observed (·) and predicted (–) serum concentrations of VRC07−523LS as a function of time in individual participants (one per plot); Group 3 (IV 20 mg/kg) is shown. The two–compartment population PK model with fully unstructured random effects variance–covariance matrix was fitted to VRC07−523LS concentrations.

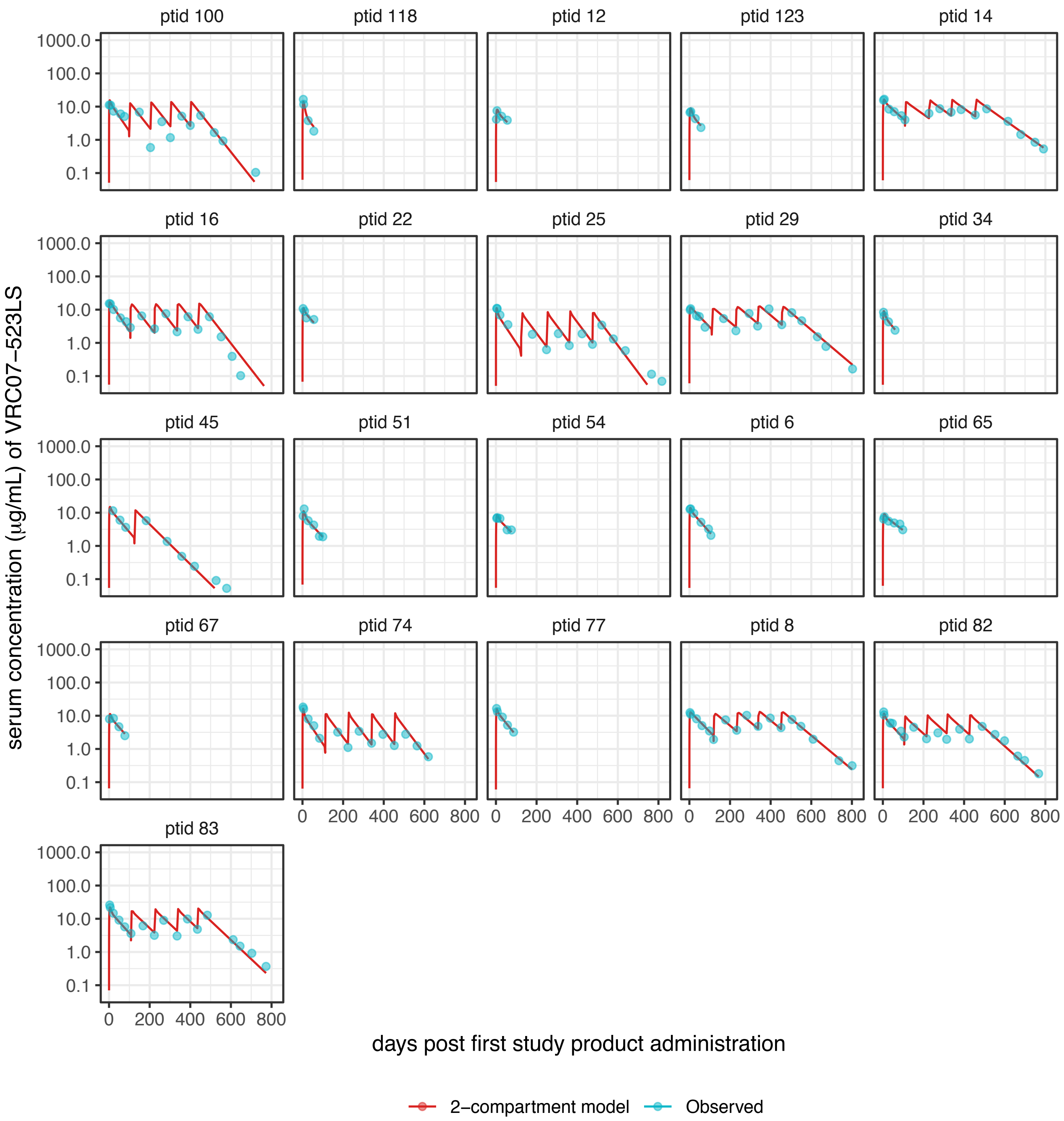

**Supplemental Figure 4D**. Observed (·) and predicted (–) serum concentrations of VRC07−523LS as a function of time in individual participants (one per plot); Group 4 (SC 2.5 mg/kg) is shown. The two-compartment population PK model with fully unstructured random effects variance-covariance matrix was fitted to VRC07−523LS concentrations.

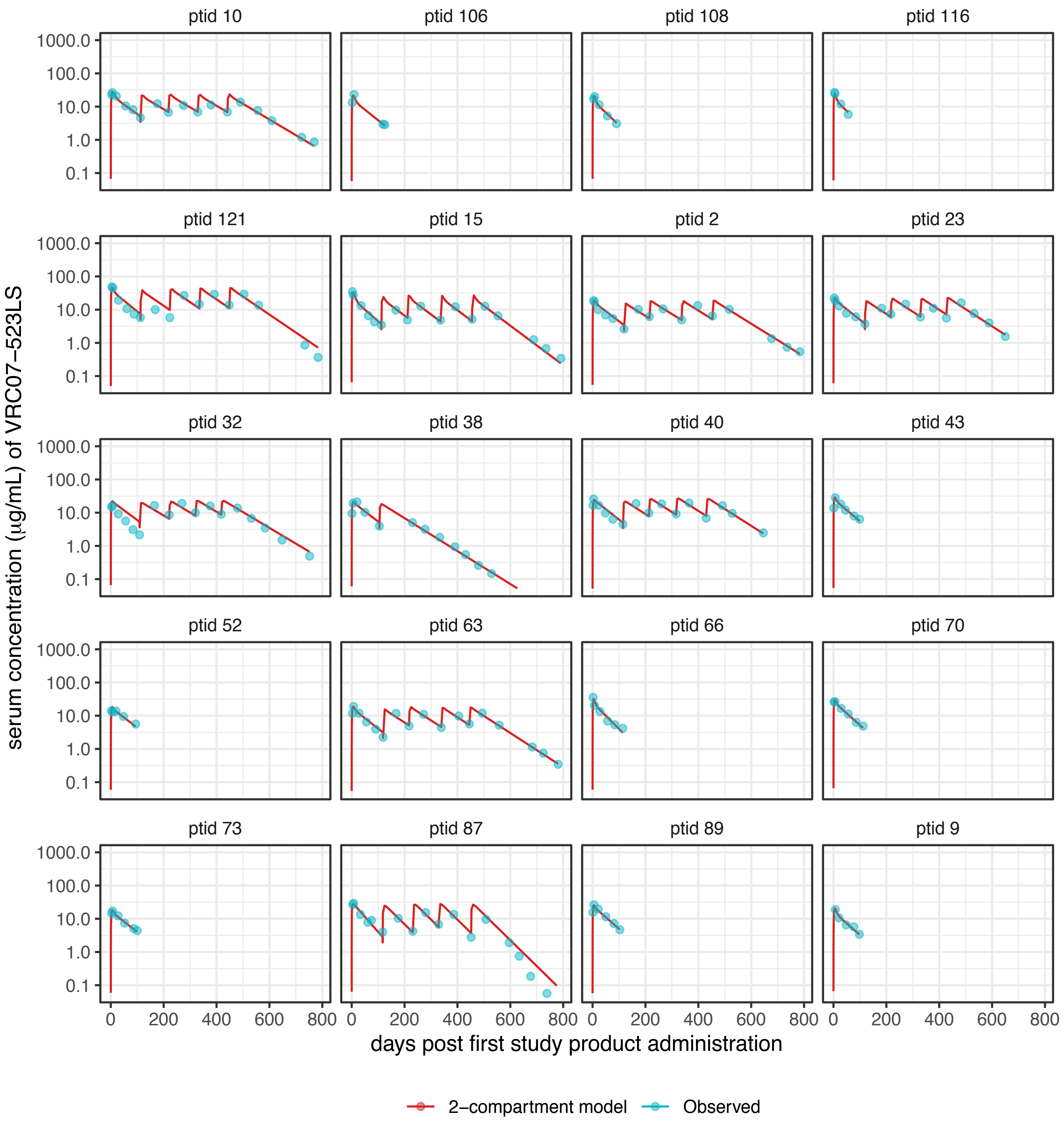

**Supplemental Figure 4E**. Observed (·) and predicted (–) serum concentrations of VRC07–523LS as a function of time in individual participants (one per plot); Group 5 (SC 5 mg/kg) is shown. The two–compartment population PK model with fully unstructured random effects variance–covariance matrix was fitted to VRC07–523LS concentrations.

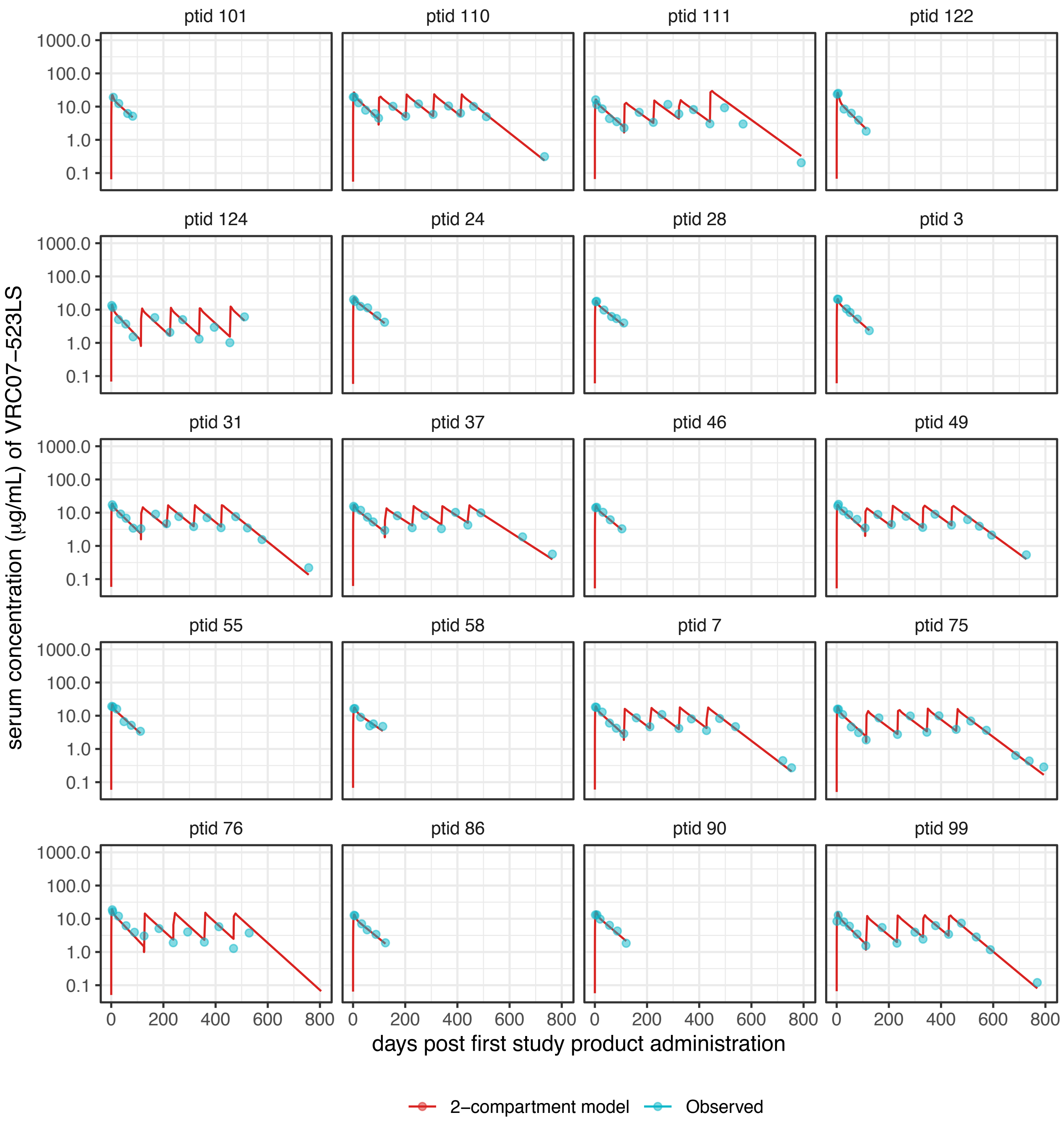

**Supplemental Figure 4F**. Observed (·) and predicted (–) serum concentrations of VRC07–523LS as a function of time in individual participants (one per plot); Group 6 (IM 2.5 mg/kg) is shown. The two–compartment population PK model with fully unstructured random effects variance–covariance matrix was fitted to VRC07–523LS concentrations.

| Parameter | Units | Estimate | SE | %RSE |
|---|---|---|---|---|
| Bioavailability for SC administration ($F_{SC}$) | - | 0.40 | 1.3e-03 | 0.32 |
| Bioavailability for IM administration ($F_{IM}$) | - | 0.57 | 0.03 | 4.67 |
| Absorption rate constant for SC administration ($Ka_{SC}$) | 1/day | 0.30 | 0.01 | 4.92 |
| Absorption rate constant for IM administration ($Ka_{IM}$) | 1/day | 0.39 | 0.02 | 4.04 |
| ELISA elimination clearance (Cl) | L/day | 0.11 | 3.1e-03 | 2.75 |
| Increase in elimination clearance from BAMA relative to ELISA ($\beta_{Cl}$) | L/day | 0.04 | 1.2e-03 | 3.21 |
| ELISA central compartment volume (V1) | L | 2.72 | 0.12 | 4.25 |
| Increase in central compartment volume from BAMA relative to ELISA ($\beta_{V1}$) | L | 0.46 | 0.03 | 6.04 |
| ELISA intercompartmental clearance (Q) | L/day | 0.49 | 0.03 | 6.68 |
| Increase in intercompartmental clearance volume from BAMA relative to ELISA ($\beta_{Q}$) | L/day | 0.69 | 0.04 | 6.08 |
| ELISA peripheral compartment volume (V2) | L | 3.59 | 0.12 | 3.35 |
| Increase in peripheral compartment volume from BAMA relative to ELISA ($\beta_{V2}$) | L | 0.15 | 2.9e-03 | 1.93 |
| Standard deviation for random effect Cl | - | 0.28 | 0.02 | 6.83 |
| Standard deviation for random effect V1 | - | 0.33 | 0.03 | 9.72 |
| Standard deviation for random effect Q | - | 0.54 | 0.05 | 9.95 |
| Standard deviation for random effect V2 | - | 0.23 | 0.03 | 11.08 |
| Correlation between Q and Cl | - | 0.15 | 0.12 | 82.95 |
| Correlation between V1 and Cl | - | 0.43 | 0.10 | 22.97 |
| Correlation between V2 and Cl | - | 1.00 | 0.02 | 1.84 |
| Correlation between V1 and Q | - | -0.33 | 0.13 | 38.27 |
| Correlation between V2 and Q | - | 0.24 | 0.15 | 60.14 |
| Correlation between V2 and V1 | - | 0.39 | 0.14 | 36.54 |
| Error model (intercept) | - | 0.19 | 0.02 | 9.65 |
| Error model (slope) | - | 0.14 | 4.6e-03 | 3.21 |
| ELISA distribution half-life | day | 1.17 | 0.13 | 10.72 |
| Increase in distribution half-life from BAMA relative to ELISA ($\beta_{DHL}$) | day | 3.44 | 0.03 | 0.92 |
| ELISA elimination half-life | day | 42.43 | 0.75 | 1.77 |
| Increase in elimination half-life from BAMA relative to ELISA ($\beta_{EHL}$) | day | 0.24 | 2.2e-03 | 0.90 |

T1: IV 2.5 mg/kg VRC07-523LS
T2: IV 5 mg/kg VRC07-523LS
T3: IV 20 mg/kg VRC07-523LS
T4: SC 2.5 mg/kg VRC07-523LS
T5: SC 5 mg/kg VRC07-523LS
T6: IM 2.5 mg/kg VRC0-7523LS

**Supplemental Table 3**: Parameter estimates based on the two-compartment population PK model fitted to VRC07-523LS serum concentrations in participants who received the study product via the IV (T1-T3), SC (T4, T5), or IM (T6) route. The model assumed a linearly combined error variance and a fully unstructured random effects variance-covariance matrix. SE: standard error; %RSE: relative SE, calculated as (SE/Estimate) × 100.

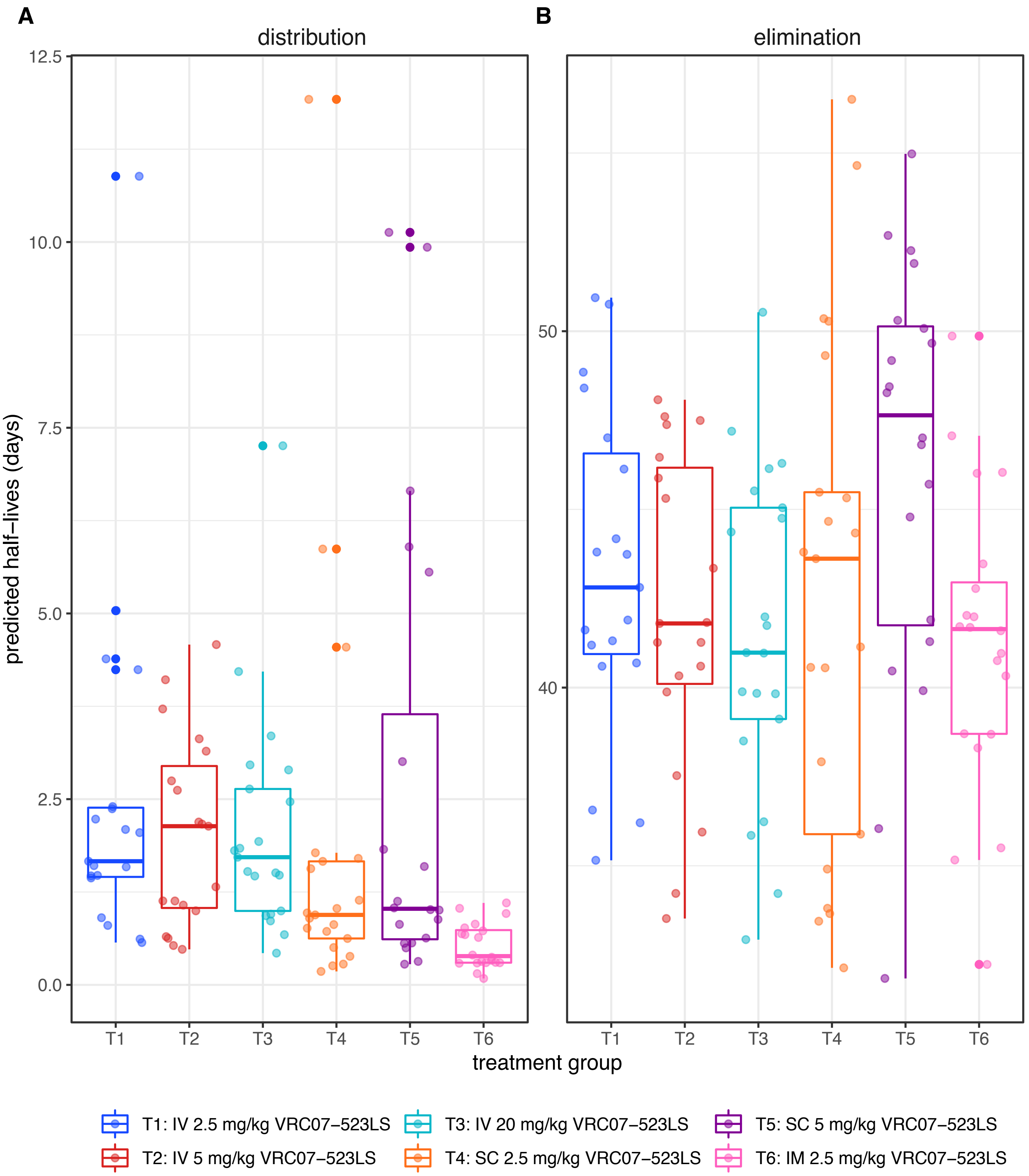

**Supplemental Figure 5.** Distribution (A) and elimination (B) half-life of VRC07-523LS. Values were predicted from the two-compartment model on a per-participant basis for each dose and route group. Box and whisker plots indicate the quartiles and X of the data set.

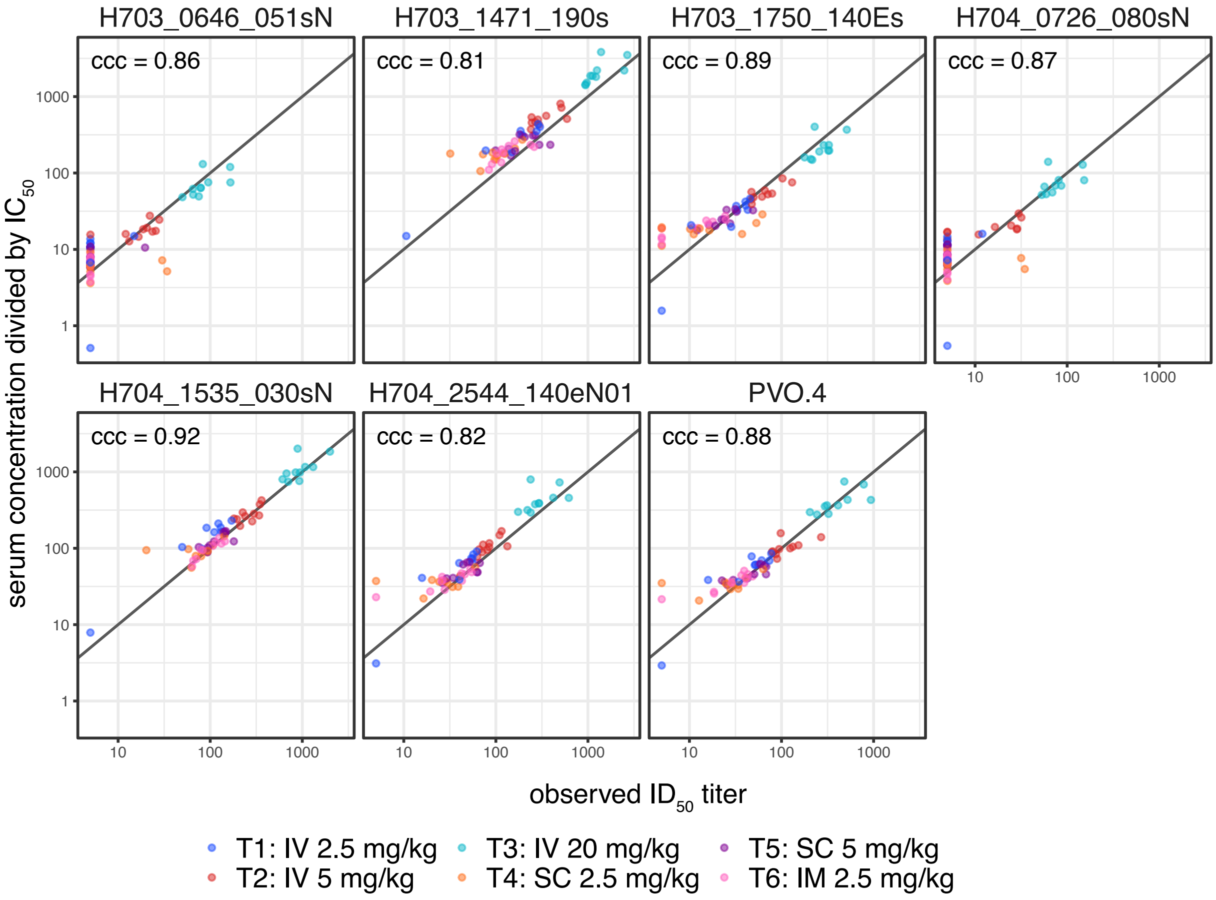

**Supplemental Figure 6.** Neutralization activity of participant serum 8 weeks following their first VRC07-523LS administration against seven HIV-1 isolates collected from incident HIV-1 acquisition events in placebo recipients in the AMP trials. $ID_{50}$ titre is shown.

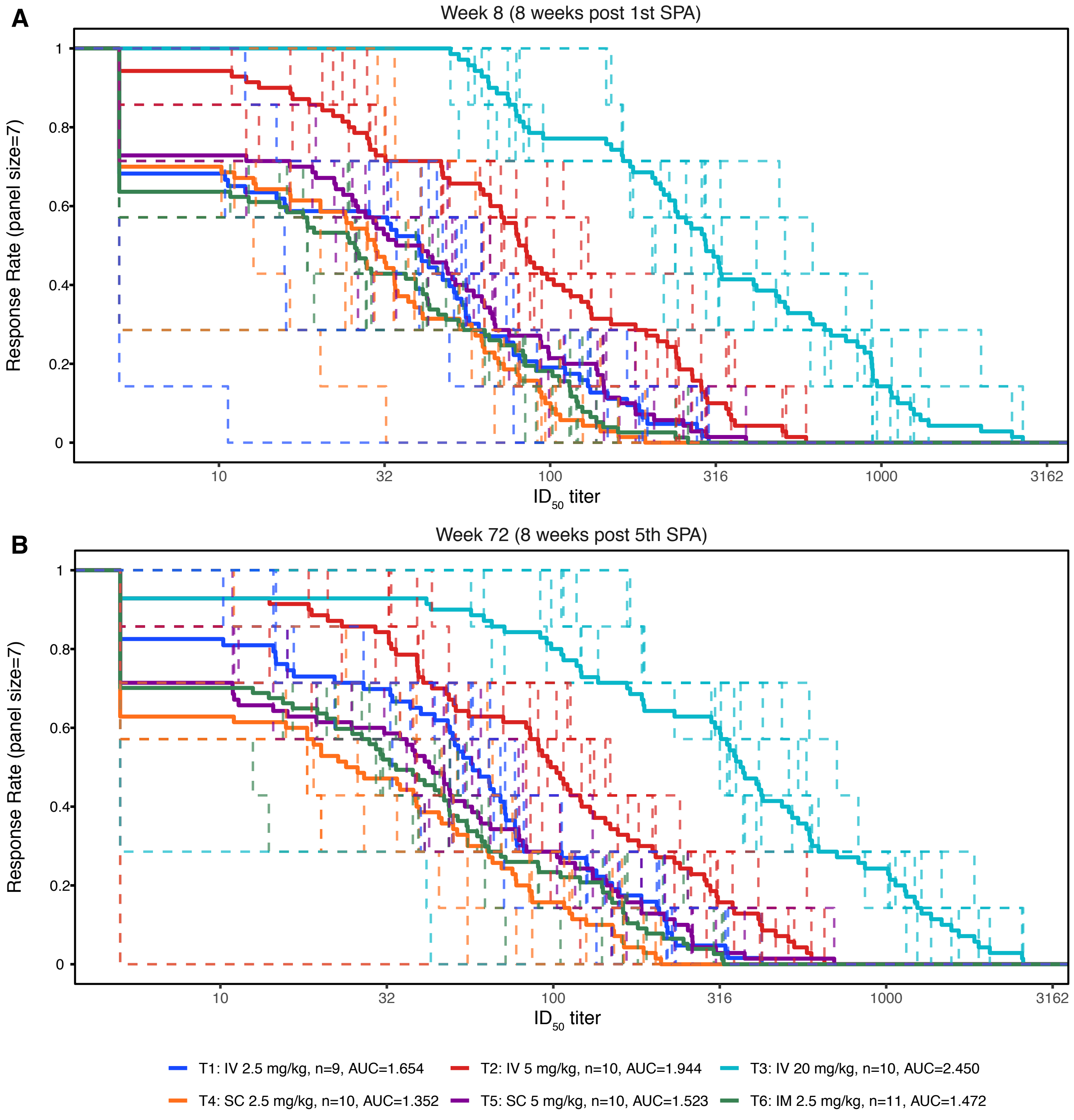

**Supplemental Figure 7.** Magnitude-breadth curves for participant serum following VRC07-523LS administration against a panel of HIV-1 isolates collected from incident HIV-1 acquisition events in placebo recipients in the AMP trials. $ID_{50}$ titre is shown at Week 8 (**A**) and Week 72 (**B**) following first study product administration.
